# Comprehensive characterization of early-stage Non-Small Cell Lung cancers with multi-modal data integration

**DOI:** 10.1101/2025.11.10.25339890

**Authors:** Daniel Schulz, Marie Morfouace, Bruno Palau Fernandez, Marcin Możejko, Sophie Déglise, Michelle Daniel, Nils Eling, Henoch S Hong, Stephanie Tissot, Sylvie Rusakiewicz, Timo Trefzer, Jane Merlevede, Flavia Marzetta, Natalie de Souza, Damian Wójtowicz, Ewa Szczurek, Benjamin Besse, Bernd Bodenmiller

**Affiliations:** Department of Quantitative Biomedicine, University of Zurich, Zurich, Switzerland; Institute of Molecular Health Sciences, ETH Zurich, Zurich, Switzerland; Institut Gustave Roussy, Villejuif, France; EORTC HQ, Avenue E. Mounier 83/11, 1200, Brussels, Belgium; Faculty of Mathematics, Informatics and Mechanics, University of Warsaw, Poland; the healthcare organization of Merck KGaA, Darmstadt, Germany; Department of Oncology, Centre Hospitalier Universitaire Vaudois, Lausanne, Switzerland; Ludwig Institute for Cancer Research, Lausanne branch, Lausanne, Switzerland; Immune Landscape Laboratory, Centre Thérapies Expérimentales (CTE), Centre Hospitalier Universitaire Vaudois, Lausanne, Switzerland; Research & Early Development Oncology, Bayer AG, Berlin, Germany; Institut Curie, PSL University, Paris, France; Institut National de la Santé et de la Recherche Médicale (INSERM), U1331, Paris, France; Mines Paris, PSL University, CBIO - Centre for Computational Biology, Paris, France; Vital-IT group, SIB Swiss Institute of Bioinformatics, Lausanne, Switzerland; Institute of AI for Health, Helmholtz Zentrum München, Germany

## Abstract

Non-small cell lung cancer (NSCLC) is one of the leading causes of cancer-related death despite the availability of therapies targeting the tumor microenvironment (TME) and/or the tumor. Immunotherapy and novel targeted therapy have improved the management of NSCLC, but the heterogeneity of the disease still hampers treatment efficacy as well as further therapeutic advances. Here, we used spatial single-cell data complemented with bulk RNAseq and whole exome sequencing to investigate the TME of 192 resected, mostly early-stage NSCLC tumors and to characterize associations of genetic, clinical and lifestyle factors with cellular and histological properties of the TME. We found, that the TME of squamous cell carcinoma (LUSC) harbored stronger signs of inflammation and immune exhaustion than adenocarcinoma (LUAD), but interestingly that elevated PD-L1 expression was associated with inflammation and markers of lymphocyte activation/exhaustion only in LUAD. Smoking correlated with T cell infiltration and TP53 mutation in LUAD, and TP53 mutation was associated with proliferation of tumor cells. In both histologies, naïve and BCL2^+^ T cells decreased with higher clinical stages. *EGFR*-driven tumors showed fewer proliferating, activated and exhausted T cells, but more CD4 T cells and HLA-DR+ tumor cells, than tumors with wild type EGFR. Multi-modal integration of our four data types showed that most variation in our cohort was along the axes of histology, smoking, *TP53* mutation, and inflammation. Our integration identified histology-specific prognostic signatures, with a LUSC-enriched profile of proliferation, inflammation, and smoking associated with poor prognosis in LUAD. In summary, we provide a rich, multimodal, single-cell characterization of a large NSCLC cohort as a resource and suggest that future investigations of biomarkers for ICI in NSCLC will benefit from stratification by histology.

## Introduction

Lung cancer is a heterogenous disease and the leading global cause of cancer-related death^1^. Non-Small-Cell Lung Cancer (NSCLC) is comprised of several histological subtypes, with adenocarcinoma (LUAD) accounting for approximately 40% and squamous cell carcinoma (LUSC) for 20-30% of cases^2^. Currently, surgical resection with curative intent is the primary treatment for early-stage lung cancer. Moreover, for non-oncogene-addicted NSCLC, immune checkpoint inhibitors (ICIs) are standard of care for almost all stages, alone or with chemotherapy, based on PD-L1 expression^3–5^.

With the arrival of neoadjuvant and adjuvant ICI in early-stage NSCLC, a better understanding is needed of which patients will respond to therapy, including the identification of biomarkers to identify these patients and to monitor their response. Single readouts such as PD-L1^6^, tumor mutational burden^7^ (TMB) or tertiary lymphoid structures^8–10^ (TLS) have been demonstrated as putative predictive or prognostic biomarkers in NSCLC, with various reproducibility. Discovery of more complex biomarkers is also in progress, with the increased use and integration of omics technologies to characterize NSCLC^11–13^. However, despite the many known differences between LUAD and LUSC, biomarker evaluation often fails to account for these distinctions.

LUAD and LUSC differ in various aspects ranging from their driver mutations^11,12^ to markedly different tumor microenvironments (TMEs)^13–15^. LUSC has been shown to contain more PD-L1 positive cells^16^, more cytotoxic and exhausted CD8 T cells^14^ and more tumor associated neutrophils (TANs)^13,15^. Furthermore, different spatial niches have been identified in samples with high neoantigen burden between LUAD and LUSC^13^. In addition, the TME also shows strong interactions with the tumor mutational landscape, and these interactions play a role in tumor initiation, progression and resistance to therapy^17,18^. For instance, previous studies have found a less inflamed TME in *EGFR*-mutated tumors^19,20^, higher PD-L1 expression in *KRAS-*mutated tumors^20^ and a more inflamed TME and PD-L1 expression in *TP53-*mutated tumors^21,22^. Interactions are also found with lifestyle factors: *TP53* mutation is more prevalent in LUSC, the dominant histological subtype associated with smoking^23^. Smoking in turn, the leading risk factor for NSCLC, is typically linked to pro-inflammatory immune cell states^24,25^. The path to improved ICI biomarkers will require a comprehensive understanding of the interplay between genetic mutations, key lifestyle factors and histology in shaping the TME. Here, we analyzed samples from 192 early-stage NSCLC patients with a multi-omics pipeline from the pan-European IMMUcan consortium^26,27^. This pipeline was used to generate imaging data with Imaging Mass Cytometry (IMC) and multiplexed immunofluorescence (mIF)^27^, hematoxylin and eosin (H&E) stains, bulk RNAseq and whole exome sequencing (WES) data. We performed detailed characterizations of clinical parameters, biomarkers, and driver mutations across histology. We confirm previously identified differences in the TMEs of LUAD and LUSC, show how in LUAD but not LUSC the immune compartment changes along with PD-L1 levels, and reveal how different driver gene and *TP53* mutations impact the TME across histologies. Our data provides a rich multi-modal resource characterizing a large early-stage NSCLC patient cohort, with a focus on the differences between the LUAD and LUSC TME and may contribute to histology-based treatment decisions in NSCLC in the future.

## Results

### Clinical and cellular characterization of the patient cohort

To deeply characterize the TME of early-stage NSCLC, we applied the multi-omics pipeline of IMMUcan^26,27^ to a clinically characterized cohort of 183 stage I-IIIA and 9 stage 4 NSCLC patients. Our cohort consisted of FFPE samples from surgical resections of primary tumors collected under the SPECTA protocol between 2014 and 2019 (**Figure 1, Methods, NCT02834884**). Median age at diagnosis was 66 (18-85) and the male to female ratio was 1.4. The majority of patient samples had a histology of LUAD (n=125, 65%), followed by LUSC (n=41, 21%). Almost all patients received curative intent surgery (n=191, 99.5%). In terms of peri-operative treatment, 14 patients received neo-adjuvant (7.3%) and 73 patients received adjuvant (38%) chemotherapy (**Supplementary Table 1**). No ICIs were given. In the metastatic settings (n=39, 20.3%), patients received chemotherapy or tyrosine kinase inhibitors if applicable. As expected, the majority of patients (n=127, 66.1%) reported past or current smoking habits (**Figure 1A**). Median follow-up was 34 months and 150 (78.1%) patients were alive at the time of database lock. The final dataset consisted of 117 samples for which all modalities were available, 60 for which all imaging modalities, WES and clinical data were available, and 15 for which a mix of the different imaging modalities, WES and clinical data were available (**Figure 1B**).

**Figure 1:**
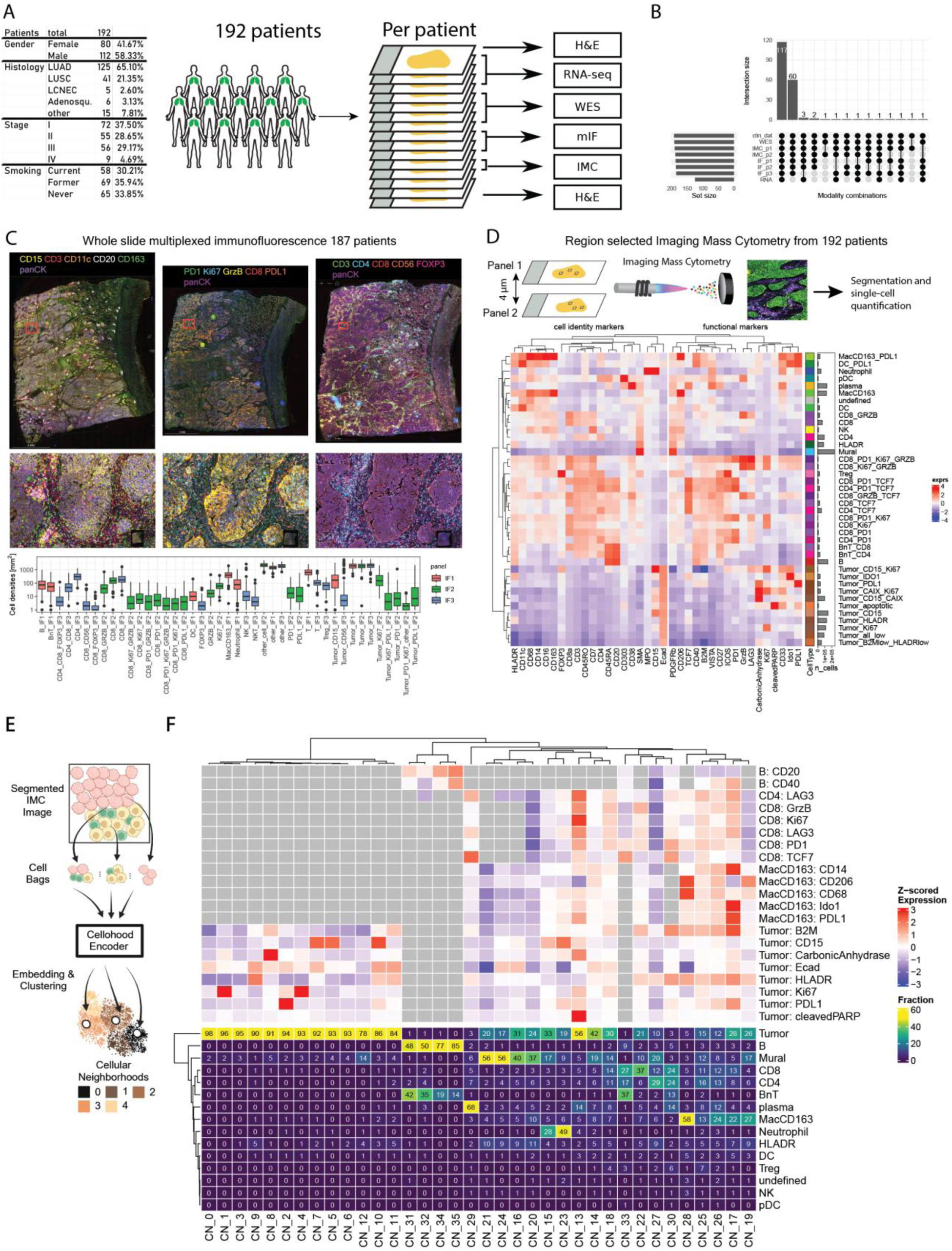
Study overview and single cell description. **(A)** Key features of the NSCLC patient cohort. **(B)** Overview of data modalities collected across the cohort. **(C)** Example images from 1 patient taken with mIF panels 1-3 (top, left to right) and summary of cell type densities in tumor areas from all mIF panels across the whole cohort (bottom). **(D)** Sketch of IMC data generation from consecutive sections using IMC panels one and two (top). Heatmap depicting the z-scaled mean expression of markers from IMC1 cell types and subtypes. Markers are split into cell type specific and functional markers. Bars indicate the abundance of cell subtypes across the cohort. **(E)** Sketch of spatial cluster generation as proposed by our companion paper Mozejko et al. Each cell gets assigned to a latent space based on their immediate cellular neighborhood and the latent space is then clustered to retrieve groups of similar neighborhoods (**Methods**). **(F)** Heatmap depicting the fraction of cell types in each cellular neighborhood (bottom) and the z-scaled mean marker expression of cell type relevant markers in each cellular neighborhood for IMC1 (top). Mean marker expressions are shown for cell type relevant markers and only if the cell type is made up at least 2 % or more of the spatial cluster.

To characterize the TME spatially at single-cell resolution, we generated multiplexed immunofluorescence (mIF) and imaging mass cytometry (IMC) data focusing on immune cell markers, as well as RNA-seq and whole-exome sequencing (WES) data, all from formalin-fixed paraffin-embedded tissue sections (**Figure 1A, Methods**). We generated mIF data with three 7-plex panels, measuring close to 500 million cells using our recently developed workflow (**Figure 1C, Methods, Supplementary Table 2**)^27^. Tumor cells were the most abundant cell type, and we observed large variation in immune cell densities, indicating heterogeneity in inflammation and infiltration between patients (**Figure 1C**).

For detailed analysis of immune-dense regions within the tumors, we utilized two complementary IMC panels focusing mainly on the detection of immune and stromal cells (e.g. lymphocytes, myeloid cells, and fibroblasts) and the expression of functional markers on these cells (**Supplementary Table 2**). A custom process for IF-co-staining and region of interest (ROI) selection was applied to select a total tissue area of ∼1 mm^2^ per patient, split into 3 ROIs focusing on regions containing tumor, immune and stromal cells, as we have previously described,^27^. This resulted in approximately 1.5 million segmented cells for IMC panel one (IMC1) (**Methods**) and 1.6 million cells for IMC panel two (IMC2). Cell type classification was performed using an annotated data set for IMC1^27^ and for IMC2 (**Supplementary Figure 1A, B, Methods**). Tumor cells accounted for 44% of all cells in IMC1 followed by myeloid cells (17%), T cells (15%), and mural cells (13%). IMC1 contained numerous markers also used to in mIF and to identify cell phenotypes in IMC that were comparable with those in the mIF data, we applied thresholds to identify marker positive cells in the data from IMC1 (**Methods**). We defined different tumor cell subtypes based on PD-L1, IDO1, Ki67, B2M, HLADR, CAIX and PARP, subtypes of macrophages and DCs based on PD-L1, and numerous CD8 and CD4 T cells subtypes based on GrzB, Ki67, PD1, and TCF7 (**Figure 1D**). IMC2 contained different markers to identify different functional states of the same and other cell types and contained a number of different T cell markers (PD1, Tim3, CXCL13, CD134, CD137, GITR, NKG2A, EOMES, GATA3, BCL2, Ki67) which we used to sub-cluster T cells (**Supplementary Figure 1C, Methods**).

The spatial arrangement of the TME can be used to stratify patients and predict treatment response^28–31^. To identify differences in the spatial arrangement of cells in our data, we made use of a novel spatial clustering method called “Cellohood” introduced in our companion paper (SUBMITTED) (**Figure 1E; Methods).** Cellohood is a permutation-invariant transformer autoencoder trained to encode latent representations for sets of proximal cells. Clustering the latent space reveals similar sets of proximal cells based on arrangement and marker expression. Cellohood for IMC1 identified neighborhoods of predominantly pure tumor cells that differed by the expression profiles of the tumor cells (CN_0-CN_12), neighborhoods dominated by mural cells and tumor cells (CN_16, 20, 21, 24), neutrophil-tumor mixed neighborhoods (CN_15, 23), macrophage rich neighborhood (CN_28), a plasma cell enriched neighborhood (CN_29) and variable neighborhoods of lymphocyte and B cell enrichment reflecting different states of B cell patches and TLS (CN_31, 32, 34, 35). Cellohood also identified multiple immune-tumor mixed neighborhoods with variable amounts of exhausted T cells, PD-L1^+^ macrophages, and T cells (e.g., CN_17, 25, 26, 28). In data from IMC2, Cellohood detected numerous different tumor neighborhoods (CN_0-CN_10) and tumor immune mixed neighborhoods (CN_11-CN_24) **(Supplementary Figure 1D)**.

We selected non-overlapping ROIs from consecutive sections for IMC1 and IMC2 to maximize the profiled area per patient, but still observed Pearson correlations above 0.6 for lymphocyte and tumor cell densities. Additionally, neutrophils from IMC1 were strongly correlated with PMN-MDSCs from IMC2 (**Supplementary Figure 2A**); given the marker differences in the two panels this indicates that most neutrophils in the tumors were of polymorphonuclear (**Supplementary Figure 2B**) and of tumor promoting nature, expressing MMP9, Arg1 and CXCL8 (**Supplementary Figure 1C**). The IMC data was generated from ∼1 mm^2^ per panel, often corresponding to about 1% of the tissue area profiled with mIF. To investigate how well the ROI-based IMC data reflected the whole-slide mIF data, we compared the area normalized cell densities from IMC and mIF (**Supplementary Figure 2C**). The strongest correlations for matched cell types were again observed for lymphocytes, neutrophils, and tumor cells, medium correlations between Tregs, NK T cells, macrophages and no correlations above 0.4 were found for cell types such as DCs. Overall, we observed a mean Pearson correlation of 0.55 for matched cell types between mIF and IMC. In line with our previous analysis^27^, these results suggested that for most cell types, the regions acquired with IMC reflect whole slide results from the mIF data.

In conclusion, we have generated one of the largest multi-modal dataset for early-stage NSCLC, consisting of whole slide mIF with comparable region-based IMC data, complemented with WES and RNAseq data. We used a novel spatial clustering algorithm to detect cellular neighborhoods and collected clinical and patient characteristics, to fully characterize the early stages of NSCLC.

### Impact of histology, stage and smoking status on the TME in NSCLC

The heterogeneity of NSCLC presents numerous challenges for defining effective biomarkers. Both the composition and architecture of the TME, for instance, are influenced by histology^11,32^, smoking^33,34^, and clinical stage^35^. There are so far only few systematic comparisons of spatially resolved single cell data between samples with different clinical characteristics^13,15^. We therefore compared the cellular composition of the TME across the two major histologies LUAD (n = 121) and LUSC (n = 41), stage, and smoking status.

LUSC was enriched for proliferating T and tumor cells, PD-L1^+^ tumor cells, tumor associated neutrophils (TANs, classified as Neutrophils in IMC1 and PMN-MDSCs in IMC2), fibroblasts, effector (CD8GrzB^+^Ki67^+^) and exhausted CD8 T cells (PD1^+^GrzB^+^), and was depleted for subtypes of macrophages and tumor cells expressing IDO1 (**Figure 2A**). Consistent with this, at the level of spatial communities LUSC was enriched for PD-L1^+^ clustered tumor cells (CN_2_P1), neutrophil-tumor mixing (CN_23_P1), clustered Ki67^+^ tumor cells (CN_4_P1), and tumor-fibroblast mixing (CN_12_P2) (**Figure 2B, C**). Further, since the spatial clusters were based on cellular marker expression as opposed to cell types, we observed differences between LUAD and LUSC that were not apparent via cell type abundances. Specifically, LUSC was enriched for clusters of EGFR^+^ tumor cells (CN_6_P2) and was depleted for clustered CD73^+^ tumor cells (CN_9_P2) and clustered CD15^+^ tumor cells (CN_5_P2, CN_7_P2, CN_10_P2) (**Figure 2B, D**). Together, these results show that the TMEs of the two major histologies of NSCLC are substantially different and suggest that they should be characterized separately to identify features relevant for the response to immunotherapy.

**Figure 2:**
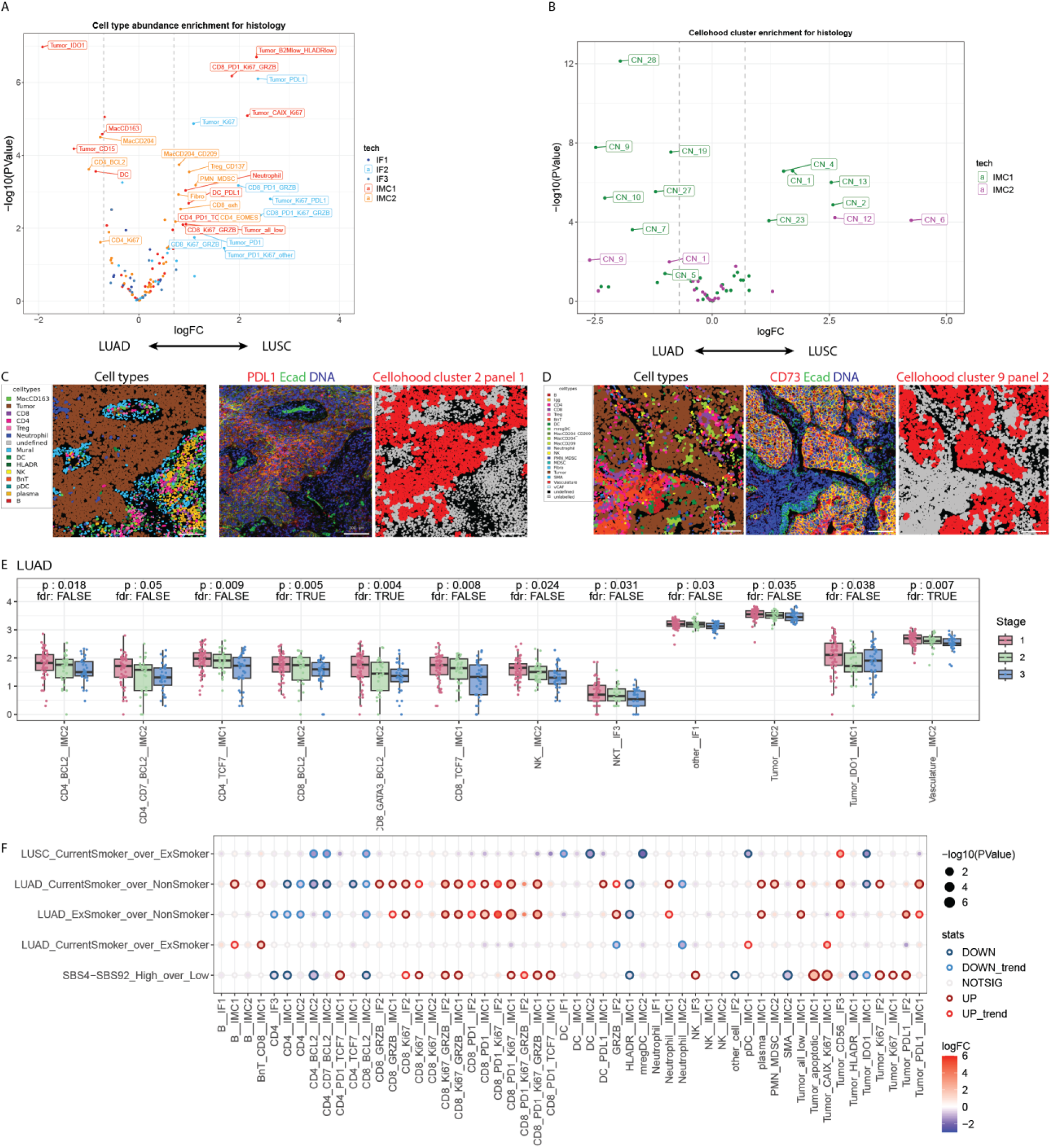
Image-feature based description of clinical characteristics. **(A)** Scatter plot showing the log fold change (X-axis) and significance (Y-axis) comparing abundance of abundance of cell types between LUSC and LUAD. Imaging modalities (IF1-3, IMC 1-2) are color coded and only cell types with FDR < 0.1 and an absolute fold change larger 0.75 are labeled. **(B)** Plot as in A but for differential abundance of spatial clusters from Cellohood. **(C,D)** Example images showing the position of classified cell types (left), the expression of the indicated markers (middle) and the position of cells from the indicated spatial clusters in red (right). **(E)** Boxplots showing differences in cell type densities across clinical stage. Only results from models with p < 0.05 are shown and FDR < 0.1 is indicated with true/false. **(F)** Bubble plot indicating the differential abundances of cell types across smoking categories separately for LUAD and LUSC and for patients with high and low smoking signatures in the WES data (bottom row). The size of each point corresponds to the p-value, the color to the log fold change and points are encircled to reflect statistical test and direction of effect (dark red: up regulation and FDR < 0.1; red: up regulation and p-value < 0.05; dark blue: down regulation and FDR <0.1; light blue: down regulation and p-value < 0.05). Cell types for which at least one test (column) was significant are shown and FDR correction was performed within each categorical test (row).

Clinical stage is also a key prognostic factor for NSCLC^36^ but whether the TME changes with stage is not well described. In our cohort, with a median follow-up time of 34 months and most patients of relatively early stage since they had been selected for surgery, we observed no difference in overall survival (OS) between stages I, II and III. However, stage IV remained a poor prognostic factor, even with the short follow-up time (**Supplementary Figure 3A**). In LUAD, we observed a decrease in anti-apoptotic (BCL2^+^), naïve (TCF7^+^) CD4 and CD8 T cells, and NK cells with increasing stage (**Figure 2E, Methods**). We could largely corroborate this finding, together with a general decrease in immune cells, in deconvolved RNAseq data from a TCGA cohort (**Supplementary Figure 3B, Methods**). In LUSC we observed a decrease in B cells, CD8 T cell subtypes, DCs, and plasma cells with increasing stage (**Supplementary Figure 3C**). In both histologies, the density of tumor cells expressing IDO1, a marker of immunosuppressive function typically upregulated upon inflammation, decreased with increasing stage.

Finally, we investigated changes in the TME associated with tobacco use. Smoking is a poor prognostic factor in NSCLC^37^ and was associated with poor OS in our cohort using univariate Cox models (Smokers vs never-smokers: HR = 4.8, p-value = 0.034; Ex-smoker vs never-smokers: HR = 2.4, p-value = 0.31). We now used our mIF and IMC data to compare cell type densities between smokers, former smokers who ceased smoking for at least 1 year, and never-smokers. In LUSC, current smokers contained fewer anti-apoptotic T cells (BCL2^+^), fewer DCs and fewer IDO1^+^ tumor cells compared to former smokers (**Figure 2F**); there were no never-smokers with LUSC histology. In LUAD we observed more effector (GrzB^+^, Ki67^+^) and exhausted (PD1^+^, CXCL13^+^) CD8 T cells and less anti-apoptotic T cells from smokers and former smokers compared to never-smokers (**Figure 2F).** Only few, poorly characterized spatial clusters showed different abundances for smoking status (**Supplementary Figure 3D**). In LUAD, a history of smoking was associated with *TP53* mutations (Odds ratio = 6.9 (smokers and former smokers compared to never-smokers, p-value = 0.037, Fisher test). There was also an enrichment of other smoking-related mutation signatures in current and former smokers (**Supplementary Figure 3E, Methods**) and the TME of patients stratified by high and low smoking-related mutation signatures partially resembled that of stratification by smoking history (**Figure 2F**). Despite cell type abundance differences suggesting ongoing inflammation in smokers, analysis of RNAseq data showed no upregulation of genes in inflammation-related pathways (JAK-STAT, NF-κB and TNFα) (**Supplementary Figure 3F**).

In summary, the TME of NSCLC varies strongly across histology, stage and smoking status. LUSC tumors are more inflamed and harbor more exhausted immune cells, fibroblasts and neutrophils than LUAD. With increasing clinical stage, the TME in LUAD and LUSC contains fewer T cells, specifically fewer CD8 T cells in LUSC. Smoking on the other hand is associated with increased abundances of CD8 T cells expressing GrzB and often proliferating but these changes are not accompanied by an inflammatory transcriptional signature.

### Clinical biomarkers PD-L1 and TMB are associated with different TMEs in LUAD and LUSC

We next investigated the TME of both major histological subtypes of NSCLC for associations with established clinical biomarkers to characterize microenvironmental features distinguishing high versus low TMB or PD-L1 expression. The FDA-approved biomarker PD-L1 tumor proportion score (TPS) and PD-L1 complete positive score (CPS) can predict ICI treatment response^38^. Even though in our cohort no ICIs were given, we wanted to investigate features in the TME that may be associated with these biomarkers. No IHC-based PD-L1 scoring was available for this cohort; therefore, we derived the TPS and CPS scores both with 1% cut-offs from the mIF data, which is similar in sensitivity to IHC^39^ (**Figure 3A, Methods**). In our cohort 30 and 49 percent of patients were classified as TPS and CPS > 1%, respectively and high TPS scores were enriched in LUSC and current smokers (**Figure 3B**). In univariate Cox models TPS > 1% was not associated to OS (HR = 1.4, p = 0.316) and CPS > 1% showed a trend of association with poor survival (HR = 1.9, p = 0.053). LUAD samples with TPS > 1% were characterized by increased abundances of multiple CD8 T cell types expressing higher levels of exhaustion related proteins (GrzB, PD1, Tim3, CXCL13, Ki67), macrophages and dendritic cells expressing PD-L1, based on IMC and mIF data (**Figure 3C**). Multiple spatial clusters of tumor-immune mixing from IMC1 (CN_17, CN_22, CN_25, CN_26) and IMC2 (CN_18, CN_19, CN_23) were also upregulated in LUAD samples with TPS > 1%. In LUSC, we observed increased abundances of macrophages and DCs expressing PD-L1 but no similar enrichment of spatial clusters of tumor-immune mixing CD8 with TPS > 1%. These results suggest that, in LUAD but not in LUSC, higher PD-L1 expression is accompanied by more inflammation and activated/exhausted CD8 T cells.

**Figure 3:**
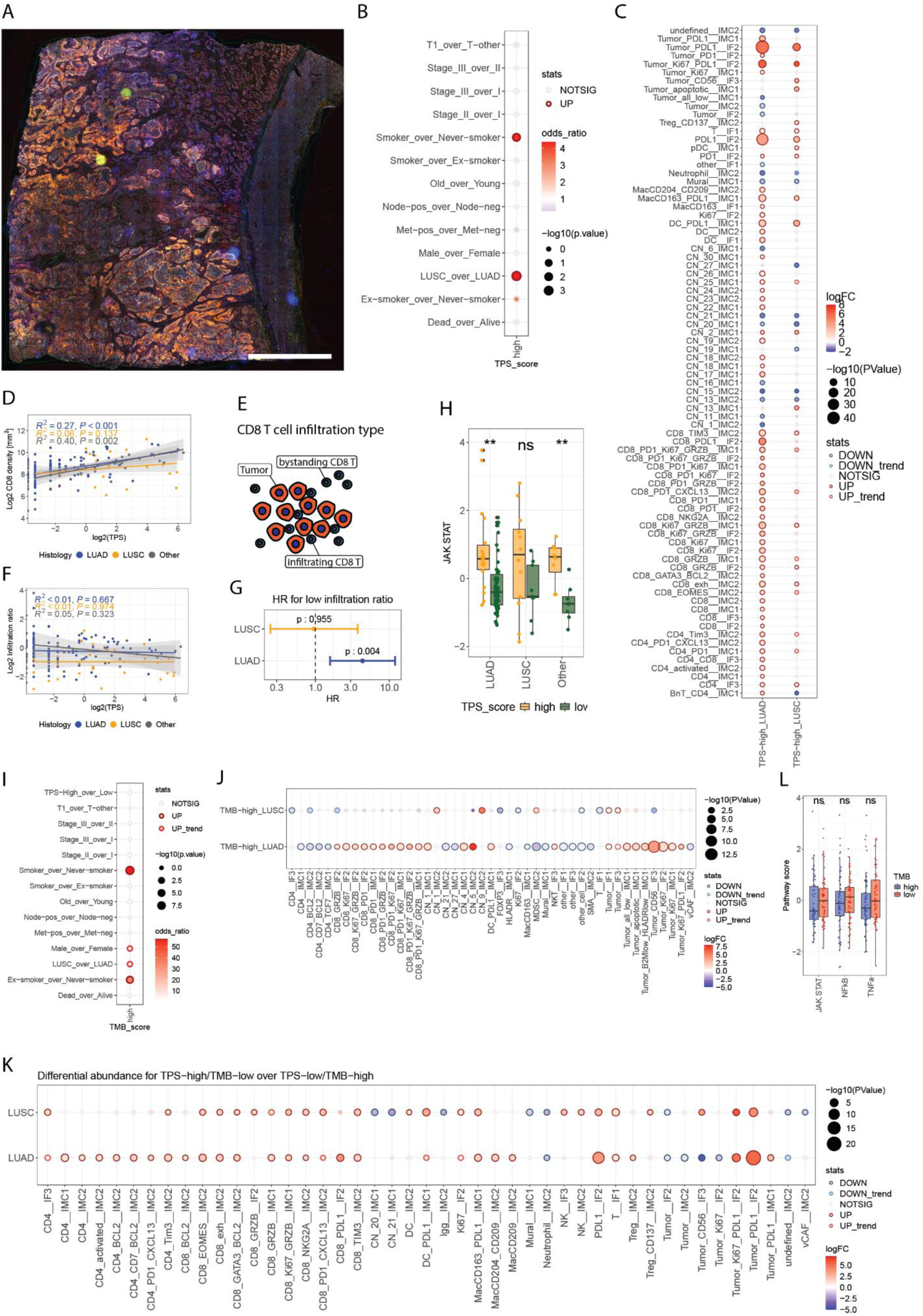
Differences in the NSCLC TME associated with clinical biomarkers. **(A)** Example image from mIF data showing the expression of PD-L1 (orange) on tumor cells. **(B)** Bubble plot indicating the enrichment of TPS > 1 % for binarized clinical parameters using Fisher tests. Points are colored by odds ratios and positive values indicate enrichment in the first string of the name (e.g. in LUSC_over_LUAD, red indicates upregulation in LUSC). The size of each point corresponds to the p-value and points with FDR < 0.1 are encircled in black. **(C)** Bubble plot indicating the differentially expressed image-based features (non-spatial and spatial) for samples with TPS > 1 % relative to samples with TPS < 1% in LUAD (left column) and LUSC (right column). Each point is sized by the p-value, colored by the log fold change and encircled according to statistical tests. **(D)** Correlation of CD8 T cell densities with TPS values for LUAD (blue), LUSC (green) and other histologies (gray). P-values from linear models are indicated on top left. **(E)** Sketch depicting the generation of an infiltration score for CD8 T cells. CD8 T cells are classified as infiltrating if they have tumor neighbors and otherwise classified as bystanding. The ratio of infiltrating over bystanding CD8 T cells indicates the proportion of infiltrating T cells. **(F)** Correlation of the infiltration ratios with TPS values for LUAD (blue), LUSC (green) and other histologies (gray). P-values from linear models are indicated on top left. **(G)** Results from Cox proportional hazards model for groups of patients with high or low infiltration ratios (median split); shown are LUSC (yellow) and LUAD (blue). P-values are indicated. **(H)** Boxplots depicting the differences in pathway activity scores (**Methods**) of JAK-STAT signaling according in LUAD, LUSC and other histologies. high, TPS > 1 %; low TPS < 1 %. P-values were calculated using t-tests. **(I)** Bubble plot indicating enrichment of TMB > 1 % for binarized clinical parameters using Fisher tests. Points are colored by odds ratios and positive values indicate enrichment in the first string of the name (e.g. LUSC_over_LUAD: red indicates upregulation in LUSC). The size of each point corresponds to the p-value and points with FDR < 0.1 are encircled in black. **(J)** Bubble plot as in (C) indicating the differentially expressed image-based features (non-spatial and spatial) for TMB-high over TMB-low samples in LUAD (top row) and LUSC (bottom row). **(K)** Bubble plot as in (C) indicating the differentially expressed image-based features (non-spatial and spatial) for TMB-high/TPS-low over TMB-low/TPS-high samples in both histologies. **(L)** Boxplots depicting the differences in pathway activity scores (Methods) of JAK-STAT signaling for TMB > 1 % across LUAD, LUSC and other histologies. P-values were calculated using t-tests. In B, C, I, J, K points are encircled to reflect statistical test and direction of effect (dark red: up regulation and FDR < 0.1; red: up regulation and p-value < 0.05; dark blue: down regulation and FDR <0.1; light blue: down regulation and p-value < 0.05). In C, J, K, FDR correction was performed per categorical test and only cell types or spatial clusters that had an FDR <0.1 for at least one categorical test are shown.

This hypothesis was further supported by a significant correlation between the densities of CD8 T cells from the mIF data with TPS values in LUAD (R^2^ = 0.27, p < 0.001) but not in LUSC (R^2^ = 0.06, p = 0.137) (**Figure 3D**). To test whether CD8 T cells engaged more frequently with tumor cells at higher TPS, we calculated an infiltration score based on the ratio of CD8 T cells with multiple versus few tumor cell neighbors CD8^28^ (**Figure 3E**) (**Methods**). We found no association between TPS and the infiltration score (**Figure 3F**). Interestingly, the dichotomized infiltration score was highly prognostic in a histology-dependent manner, with low infiltration ratios having poor prognosis in LUAD (HR = 3.5, p = 0.01) but not in LUSC (HR = 0.96, p = 0.95) (**Figure 3G**). Further, TPS >1% tumors showed a JAK-STAT gene expression signature of inflammation in LUAD but not in LUSC (**Figure 3H**).

The tumor mutational burden (TMB) is also an approved biomarker for ICI treatment of NSCLC. To understand the impact of high TMB on the TME, we compared cell densities in the imaging data (mIF and IMC) in samples with low and high TMB, defining the latter as > 10 mut/Mb^40^ derived from our WES data (**Methods**). TMB-high patients were more likely to be smokers, male, and with tumors of LUSC histology (**Figure 3I**). In LUAD, high TMB was associated with more proliferating, GrzB^+^ and PD1^+^ CD8 T cells, fewer CD4 T cell subtypes, more proliferating tumor cells, CD56 expressing tumor cells (also CN_5_IMC2), and tumor cells expressing lower levels of antigen presentation related proteins HLADR and B2M (**Figure 3J**). In LUSC, high TMB was associated with fewer CD4 T cells, enriched for CD73 expressing tumor cell clusters (CN_9_IMC2) and showed no upregulation of any CD8 T cell subtypes. While TMB high samples in LUAD were enriched for numerous proliferating CD8 T cell types and thus appeared similar to TPS-high samples, many T cell subtypes were enriched in TMB low/TPS-high compared to TMB high/TPS-low samples in LUAD and LUSC suggesting that TMB is associated with smaller T cell infiltration and activation than TPS > 1% (**Figure 3K)**. TMB-high and TPS > 1% were not associated (Fishers exact p = 0.08) (**Figure 3I**) and we found no significant association between the expression of inflammatory pathway genes and TMB scores **(Figure 3L)**.

In summary, while the TME of LUSC generally contained more PD-L1 and activated/exhausted CD8 T cells than LUAD, we observed minimal changes in the LUSC TME with varying PD-L1 or TMB levels. In LUAD however, higher levels of PD-L1^+^ cells, and to a lesser extent high TMB, were associated with higher numbers of lymphocytes expressing activation/exhaustion markers.

### B cell patches are abundant in NSCLC

Tertiary lymphoid structures (TLS) have been associated with good response to ICI^8,9,41^ and with OS in some studies^42^. Since our mIF panels did not include markers to investigate the functional state of TLS – such as CD21, CD23 and BCL6 – we used the mIF data to analyse the presence of B cell patches (**Supplementary Figure 4A, Methods**), which include but are not limited to TLS. Overall, in 58% of patient samples we identified at least 10 patches containing at least 100 B cells confirming that B cell aggregation occurs frequently in NSCLC, with up to hundreds of B cell patches in single sections and a large range between sections (**Supplementary Figure 4B**). B cell patches occurred proportionally more frequently inside of tumor compared to surrounding healthy normal tissue (**Supplementary Figure 4C**) but patches in the healthy normal tissue were significantly larger than those inside tumors (**Supplementary Figure 4D**), and contained relatively more B cells and fewer T cells (**Supplementary Figure 4E**). We found no difference in the density normalized B cell patch count between LUAD and LUSC (**Supplementary Figure 4F**). B cell patch identification is mostly a function of B cell density, and we observed very similar correlations for cell types with B cell or B cell patch densities with some larger correlation values for TCF7^+^ CD8 T and CD4 T cells (**Supplementary Figure 4G**). The density of TLS in tumors was neither associated with TPS nor TMB (**Supplementary Figure 4H**) and the density of B cell patches was not different between clinical stages (**Supplementary Figure 4I**). We observed a non-significant trend of B cell patches being associated with better OS in a univariate Cox model in LUAD (HR = 0.39, p = 0.055) and not in LUSC (**Supplementary Figure 4J**). We conclude that B cell aggregation is very abundant in NSCLC, but was not associated with PD-L1, TMB, histology or OS in our cohort.

### Driver mutations generate distinct TMEs

Oncogene addicted tumors are commonly observed within LUAD and these patients typically receive targeted therapy rather than ICI, to which response rates are known to be poor^43,44^. To assess the effect of oncogenic mutations on the TME, we used our WES data to detect genetic variants and identify oncogenic driver and tumor suppressor mutations (**Methods)**. Mutations were identified at lower, but relatively similar frequency as previously described^45^, with *TP53*, *KRAS* and *EGFR* being the most common (33%, 19% and 8%, respectively) (**Figure 4A**). *KRAS* and *EGFR* mutations were enriched in *LUAD* and *PIK3CA* and *TP53* mutations in LUSC (**Supplementary Figure 5A**). *EGFR* mutation was associated with never smokers, low TMB, and female patients, *KRAS* mutation with smoking, and *TP53* mutation in LUAD with high TMB (**Supplementary Figure 5A**). We then compared the abundances of cell types and spatial clusters (**Figure 4B**) and the expression of relevant markers per cell type (**Supplementary Figure 5B, Methods**) between oncogene mutated and wild type samples. In *EGFR-*mutated tumors, spatial cluster CN_6_IMC2 (characterized by the expression of EGFR itself) was most strongly upregulated. These tumors also contained lower abundances of different CD8 PD1^+^ cell types and PMN-MDSCs/Neutrophils, higher abundances of CD4 T cell types, and showed widespread expression and upregulation of HLA-DR on tumor cells (**Figure 4C**) (**Supplementary Figure 5C**). Spatial clusters CN_13_ IMC1 and CN_17_ IMC1, characterized by a mix of activated/exhausted tumor cells, had lower abundances in *EGFR-*mutated tumors, with lower expression of CXCL13, LAG3, PD1, and Tim3 on CD8 T cells (**Figure 4D**). In contrast, the TME of *KRAS-*mutated tumors in LUAD was almost indistinguishable from *KRAS* wild type tumors but contained more PD-L1^+^ tumor cells based on mIF but not based on region-selected IMC (**Figure 4B**).

**Figure 4:**
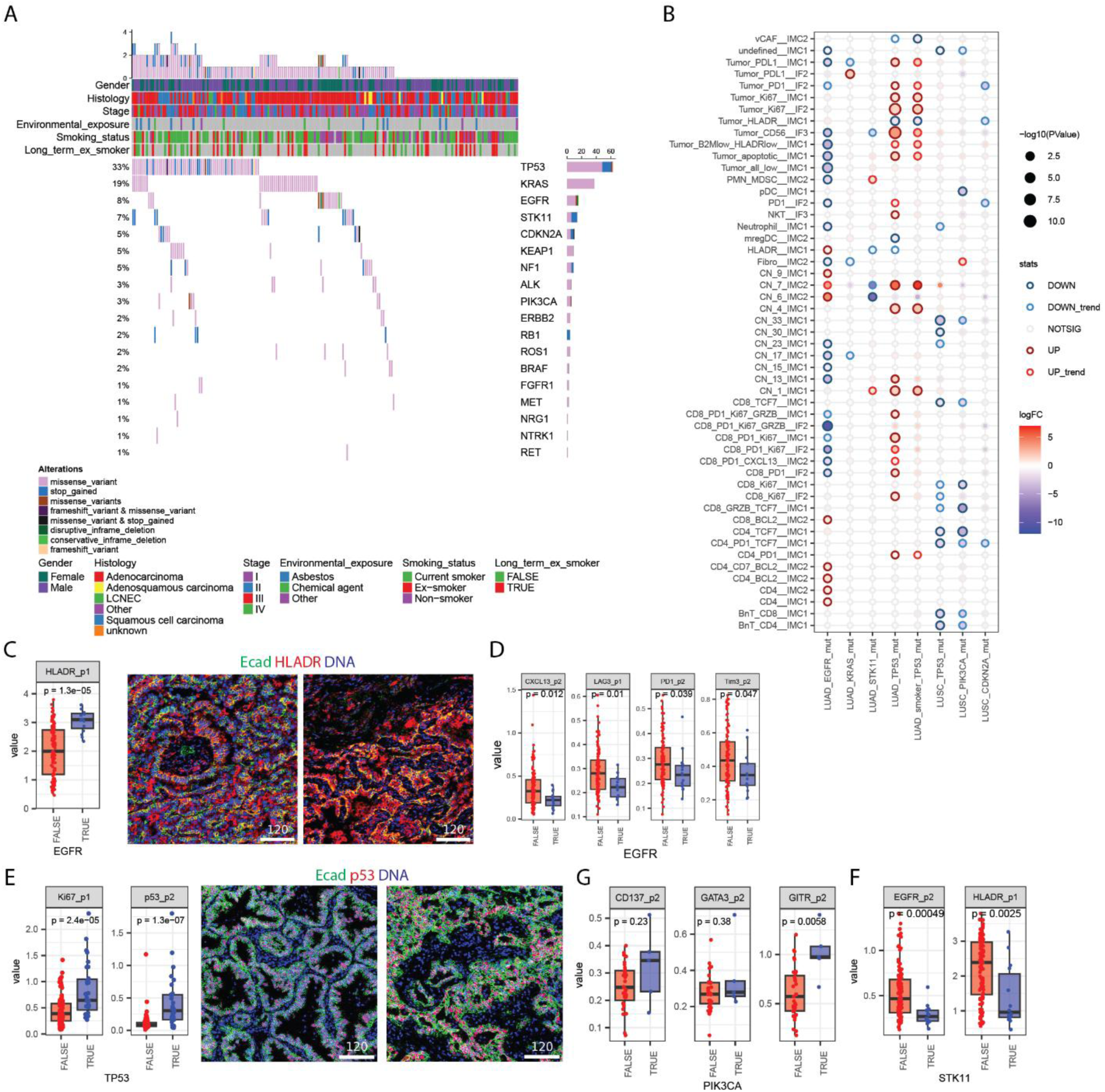
Effect of driver mutations on the NSCLC TME. **A)** Waterfall plot depicting disruptive mutations of oncogenes and tumor suppressor genes sorted by their frequency. Color coded clinical annotations and total disruptive mutation count are shown on top. **B)** Bubble plot indicating the differentially abundant cell subtypes and spatial clusters for *EGFR-*, *KRAS-*, *STK11-* and *TP53-*mutated tumors in LUAD, and *PIK3CA-* and *TP53-*mutated tumors in LUSC, in all cases compared to tumors wild type for the corresponding gene. Each point is sized by the p-value, colored by the log fold change and encircled to reflect statistical test and direction of effect (dark red: up regulation and FDR < 0.1; red: up regulation and p-value < 0.05; dark blue: down regulation and FDR <0.1; light blue: down regulation and p-value < 0.05). In C, J, K, FDR correction was performed per categorical test and only cell types or spatial clusters that had an FDR <0.1 for at least one categorical test are shown. **C)** Boxplot showing the average expression of HLA-DR on tumor cells per patient for *EGFR* mutated and wild-type groups and example images of HLA-DR expression on tumor cells (E-cadherin: green; HLA-DR: red; DNA: blue). **D)** Boxplots showing the average expression of CXCL13, LAG3, PD1 and TIM3 on CD8 T cells per patient sample for *EGFR* mutated and wild-type groups in LUAD. **E)** Boxplots showing the average expression of EGFR and HLA-DR on tumor cells per patient sample for *STK11* mutated and wild-type groups. **F)** Boxplots showing the average expression of Ki67 and TP53 on tumor cells per patient sample for *TP53* mutated and wild-type groups in LUAD and example images of TP53 expression in tumor cells (E-cadherin: green; TP53: red; DNA: blue). **G)** Boxplots showing the average expression of CD137, GATA3 and GITR Tregs per patient sample for *PIK3CA* mutated and wild-type groups in LUSC.

In *TP53* mutated LUAD tumors, we observed an upregulation of spatial cluster CN_7_IMC2, characterized by the expression of *TP53*, spatial clusters characterized by Ki67 expression on tumor cells (CN_1_IMC1, CN_4_IMC1), and Ki67^+^ tumor cells from IF2 suggesting that *TP53* mutated tumors in LUAD are more proliferative and expressed higher levels of TP53 than *TP53* wild type tumors (**Figure 4B, 4E**). We did not observe higher proliferation of tumor cells based on Ki67in *TP53-*mutated LUSC tumors, which showed generally higher proliferation based on Ki67 compared to LUAD (**Supplementary Figure 5D**). CD8 T cell types associated with immune activation/exhaustion (GrzB^+^, PD1^+^, Ki67^+^) were more frequent in *TP53-*mutated LUAD tumors (**Figure 4B**). However, TP53-mutated LUAD tumors were also enriched in smokers **(Supplementary Figure 5A)** and within smokers (n = 56) the TME of *TP53-*mutated vs non-mutated tumors was similar, suggesting that smoking, and not TP53 mutation, drives inflammation (**Figure 4B**). In LUSC, *TP53-*mutated tumors had fewer BnT cells, TCF7^+^ T cells, and spatial clusters characterized by high T cell and BnT cell frequencies (CN_30_IMC1, CN_33_IMC1). *PIK3CA*-mutated tumors were similar to *TP53*-mutated tumors, potentially because 4 of the 6 *PIK3CA*-mutated tumors were also *TP53*-mutated. However, while we did not observe a difference in the abundance of Tregs, the expression of GITR was increased in Tregs in LUSC, suggesting a more activated state of Tregs in PIK3CA*-*mutated tumors (**Figure 4G, Supplementary Figure 5E**). *STK11-*mutated tumors had lower abundances of spatial cluster CN6_IMC1 (EGFR^+^ tumor cells) and lower expression of HLA-DR on tumor cells (**Figure 4B, F**).

In summary, different driver mutations have a distinct influence on the TME and the tumor cells of both histologies of NSCLC. *TP53*-mutated LUAD tumors were characterized by higher proliferation, *EGFR*-mutated tumors contained fewer activated CD8 T cells and more CD4 T cells and expressed high levels of HLA-DR. In LUSC a population of GITR^+^ Tregs was observed in *PIK3CA*-mutated tumors.

### Multi-modal integration to link molecular, cellular and clinical variation in NSCLC

To uncover coordinated (i.e., cross-modality) and modality-specific sources of variation across our clinical, molecular and cellular data, we applied Multi-Omics Factor Analysis^46^ (MOFA), an unsupervised framework for integrative dimensionality reduction (**Methods**). MOFA identifies key patterns of variation shared across clinical, molecular, and cellular data. Each pattern, or latent factor (LF), captures coordinated changes that may reflect underlying biology or patient differences. These latent factors allow us to visualize patient relationships, explore variability, and connect molecular profiles with clinical features. Our data yielded a total of 4541 features (e.g., cell type densities, spatial patterns, mutations, marker patterns etc.) from the 4 modalities across all patients (**Figure 5A**).

**Figure 5:**
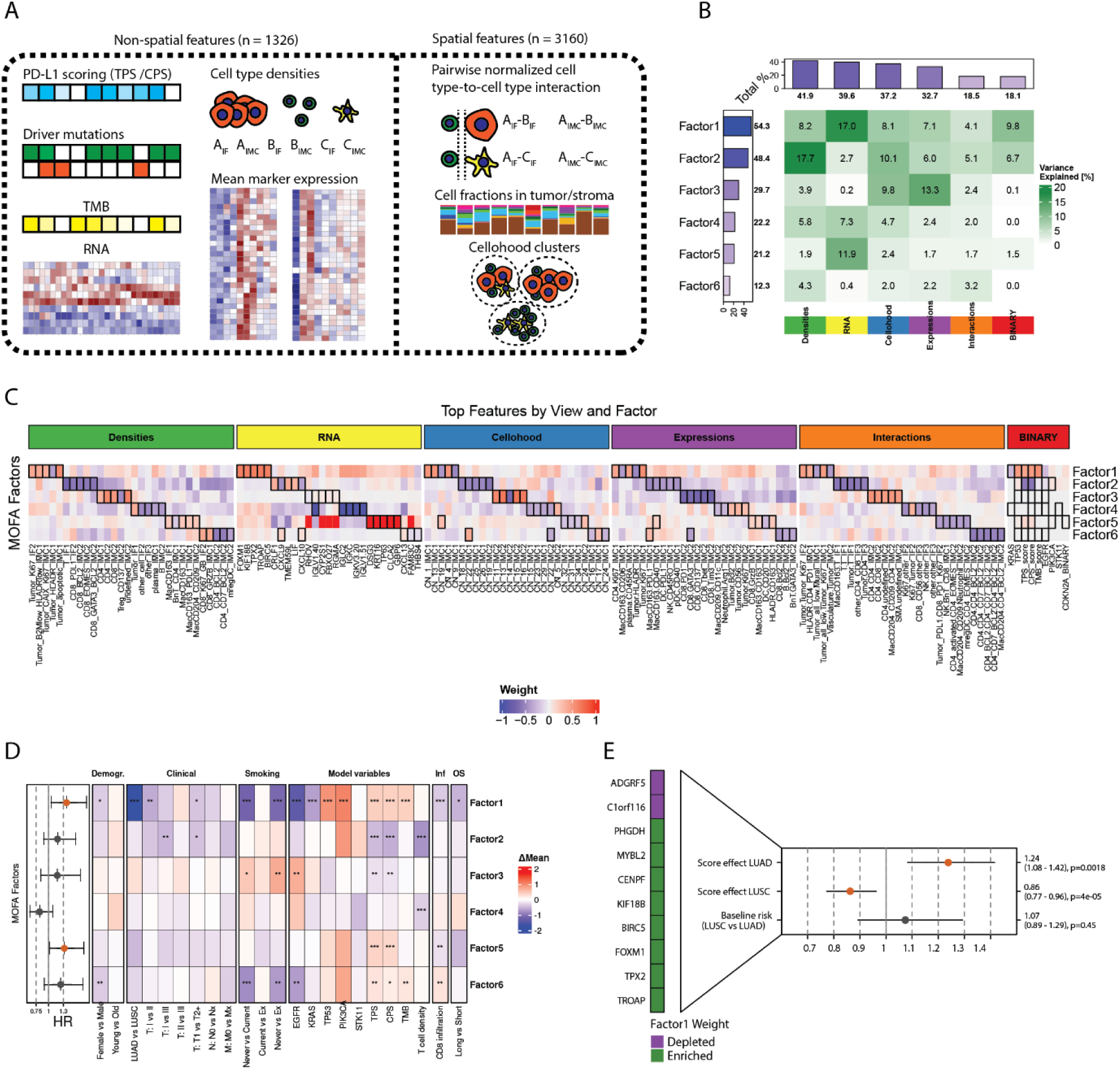
Data integration with MOFA. **(A)** Data types used as input for MOFA. **(B)** Heatmap indicating the explained variance for a given data type for each of the six latent factors. The plots show the sum of explained variance for each data type across LFs (top) and the sum of explained variance for each LF (left). **(C)** Heatmap showing distinguishing features per data type and LF. For each LF the top 5 features were selected based on absolute weights (boxes). **(D)** Heatmap depicting the association of LFs (rows) with the indicated binarized parameters (columns). The color scale indicates the difference in the mean of the factor values and significances were calculated using t-tests for categorical features or Spearman correlation tests for continuous features (TMB, TPS, CPS, T cell density, and CD8 infiltration) and were corrected for multiple-testing using Benjamini-Hochberg. Significance levels are indicated as follows: fdr < 0.001 = ***, fdr < 0.01 = ** and fdr < 0.05 = *. The plot on the left shows a Cox model comparing hazard ratios for continuous values for LF1. **(E)** The top ten genes with the highest absolute weights associated with LF1 are shown on the left. Results of a Cox model using histology and the enrichment score values for these ten genes (as interaction term) are shown. Significant HRs are shown in orange and non-significant results are shown in gray. Data from 1045 patients.

We identified 6 LFs in our dataset, several of which explained variance across multiple data types, thus reflecting coordinated changes captured in the multi-modal data (**Figure 5B**). To interpret the LFs, we plotted the weights of the top 5 most important features per data type and LF (**Figure 5C**) and used gene set enrichment analysis (GSEA) to identify varying gene sets among the LFs (**Supplementary Figure 6A, Methods**). In brief, the LFs were associated with: medium T cell infiltration, highly proliferating tumors with *TP53* mutation, high TMB and PD-L1 (LF1); cold, PD-L1 negative tumors (LF2); T helper cell enriched tumors with low activation on CD8 (LF3); cold tumors with proliferation and neuroendocrine features enriched in LCNEC histology (LF4); medium T cell densities, PD-L1 positive, differentiated, keratinizing tumors (LF5) (**Supplementary Figure 6B**); inflamed, T cell and macrophage infiltrated tumors (LF6).

We further probed the association of the LFs with demographic, clinical and molecular parameters, and found relationships consistent with these gene set-based annotations (**Figure 5D**). LF1, the factor explaining most variation in the data, was significantly associated with LUSC, smoking, *TP53* and *PIK3CA* mutation and depleted for the typical LUAD driver genes *KRAS* and *EGFR*. Further, LF1 correlated with TPS, CPS and TMB, was negatively correlated with CD8 infiltration and showed an increased hazard for OS (HR=1.36, p=0.019). However, separated by histology, LF1 was a poor prognostic factor only in LUAD (HR=1.51, p=0.05) and not in LUSC (HR=1.34, p=0.52) (**Supplementary Figure 6C**), despite its significant association with LUSC. LF5 showed similar trends as LF1 and was also associated with an increased hazard for OS (HR=1.32, p=0.039). We validated our findings with LF1 in TCGA RNA-Seq data by using genes associated with LF1 to define TCGA patient groups corresponding to those high in LF1 in our dataset (**Figure 5E**, **Methods**). This confirmed that patient groups with a gene signature corresponding to LF1 had poor prognosis in LUAD (HR=1.24, p= 0.0018) (**Figure 5E**). Interestingly, the expression of the same signature had good prognosis in LUSC in TCGA (HR=0.86, p=4.3*10^-5^) reiterating the differences between histologies.

In summary, data integration using MOFA showed that, also when simultaneously taking all measured variables into account, the strongest variation in our NSCLC cohort could be attributed to histology, smoking and *TP53* mutation. Tumors with inflammation were found across histologies but were not prognostic in this cohort. Notably, a LUSC-associated signature of inflammation and smoking was of poor prognosis when present in LUAD patients.

## Discussion

Immunotherapy is now approved for early-stage NSCLC in the peri-operative setting, but biomarkers to guide patient selection for this treatment are limited. A better understanding of the TME in NSCLC is therefore crucial. Here, we combined longitudinal clinical information, molecular profiling of the tumor, and cellular profiling of the TME, on a cohort of nearly 200 early-stage NSCLC patients. The spatial resolution of the data allowed for identification of cellular neighborhoods, associated with different clinical and molecular patterns.

Our data confirmed several known differences between the immune landscapes of LUAD and LUSC, for instance that LUSC presents fewer macrophage subtypes^47^, higher neutrophil abundances^13,15^, increased numbers of activated CD8 T cells (expressing PD1)^14^, and higher numbers of CD10^+^ or Podoplanin^+^ fibroblasts^48^. We also confirmed that LUSC tumors are more proliferative^15,49^ and express more PD-L1^16^. Although LUSC exhibited generally higher immune activation, exhaustion, and PD-L1 levels than LUAD, these features were largely independent of PD-L1 TPS. This contrasts with LUAD, where TPS >1% was linked to increased CD8 T cell infiltration and activation and exhaustion. Together, these results suggest fundamental histology-specific differences in how the tumor microenvironment senses and responds to inflammation — adaptive and inducible in LUAD and rather constitutive in LUSC. Further, TMB-high tumors were enriched for CD8 T cell subtypes in LUAD, but not in LUSC, but this infiltration effect was not as strong as in tumors with high (> 1%) TPS. In both histologies, CD4 T cells were depleted in TMB-high tumors. We did not observe a higher frequency of tumors with high TMB (>10) in LUSC versus LUAD, as previously reported in large cohorts^50^ potentially due to smaller patient numbers in our cohort or a general overestimation of the TMB due to lacking matched normal controls.

NSCLC tumors with driver genes (such as *EGFR*, *ALK* or *RET*) respond poorly to immunotherapy^44^, and in the case of EGFR, our data suggest a hypothesis for the TME underlying this effect. In particular, we observed lower numbers of exhausted CD8 T cells, tumor-immune mixing, and TANs, but more CD4 T cells, in *EGFR*-mutated tumors. Interestingly, all tumor cells in *EGFR*-mutated tumors also expressed HLA-DR; together with the higher abundance of CD4 T cells in these tumors, this indicates that CD4 T cells recognize tumor antigens via HLA-DR. This specific TME, high in CD4 T cells and lacking exhausted CD8 T cells, the major target of ICI, may at least partially explain the well-known poor response to ICI of NSCLC with EGFR mutations^44^. In *PIK3CA*-mutated tumors we observed an upregulation of GITR, a marker typically linked to Treg activity. *PIK3CA*-mutation activates PI3K thereby stimulating glycolysis and the production of lactate^51^ which can activate and stabilize Tregs^51^, suggesting that Treg targeting could be an option in those tumors. *TP53*-mutation was associated with a history of smoking, with both smokers and former smokers showing a more inflamed TME than never-smokers, despite former smokers being defined as having stopped smoking for at least one year. When investigated within smokers, however, *TP53* mutation was not associated with changes in immune compartment, suggesting that *TP53* mutation itself does not alter the TME.

The presence of TLS has been associated with favorable responses to immunotherapy in NSCLC^10,52^. Our mIF panel did not contain markers to distinguish TLS from B cell accumulations, therefore we quantified B cells patches across tumors. This analysis revealed that B cell patches of at least 100 densely packed B cells occurred frequently in NSCLC, irrespective of histology. In agreement with previous reports, the presence of B cell patches was associated with TCF7^+^ T cell densities^53^ and while B cell patches were not prognostic for OS overall in LUSC, they tended to be associated with good prognosis in LUAD.

Multi-modal integration identified latent factors describing shared variation across data modalities. Of these, two factors in particular (LF1 and LF2) explained most of the variation across our dataset. LF1 was associated with T cell exhaustion, TP53 mutation, LUSC, and smoking while LF2 was mainly associated with cold tumors. LF1 was also associated with a poor prognosis in LUAD, and we confirmed this observation in an independent TCGA cohort using a derived RNA seq-based signature. This suggests that in early-stage NSCLC, inflammation and proliferation are associated with poor survival in LUAD but not in LUSC. Our second latent factor (LF2), associated purely cold, later stage tumors, was not enriched for any histology or driver mutation and showed no effect for OS. Since we observed that the combination of *TP53* mutation, proliferation, and PD-L1 expression (i.e., LF1) is detrimental in NSCLC, we speculate that patients enriched for this signature would potentially benefit from a combinatorial treatment of chemotherapy and ICI. Similarly, patients enriched for LF6 could potentially benefit from immunotherapy-only treatment due to the presence of exhausted T cells, TCF7^+^ T cells and PDL1 expression on tumor and myeloid cells. Our observations that inflammation and tumor proliferation (LF1) are associated with poor OS are consistent with the tumor-promoting effects of inflammation^54,55^ and a study suggesting that inflammation reduction is associated with a decreased risk of lung cancer occurrence^56^.

In conclusion, we have generated a unique multi-modal dataset for NSCLC, integrating molecular and single-cell imaging data in a single resource to enable future research and methodological development. Our integrated analysis reveals that early-stage NSCLC comprises distinct immune landscapes driven by histology, oncogenic context and life-style factors. LUSC displays constitutive immune activation and T cell exhaustion, whereas LUAD shows an adaptive response linked to PD-L1 expression and TMB. A smoking-related inflammation-proliferation signature was associated with poor prognosis in LUAD but favorable outcomes in LUSC, highlighting fundamental histology-specific differences in NSCLC and in how the TME affects disease progression.

## Data Availability

All data will be available upon peer-review publication

## Acknowledgements

The IMMUcan project has received funding from the Innovative Medicines Initiative 2 Joint Undertaking under grant agreement No 821558. This Joint Undertaking receives support from the European Union’s Horizon 2020 research and innovation program and EFPIA https://IMI.europa.eu. The SPECTA Platform is supported by Alliance Healthcare. Alliance Healthcare will become Cencora.

## Author Contributions

DS, M Morfouace and B Bodenmiller designed the study. MD, SD, and DS performed IMC experiments. SR and ST performed mIF experiments. DS analyzed data with input from JM, FM, TT, and NE. DW calculated mutation signatures. BP and DS performed integration analysis. M Mozejko generated Cellohood data. M Morfouace and HSH oversaw the molecular and cellular TME profiling within IMMUcan. ES, HSH, B Besse, and B Bodenmiller gave input during project progression. DS and M Morfouace wrote manuscript with input from all authors.

## Methods

### Patient cohort

Patients were selected retrospectively from the SPECTAlung study^57^. We selected patients with a diagnosis of NSCLC at any stage, but with a surgical specimen (no biopsy) and with tumor formalin-fixed paraffin-embedded (FFPE) material available. The majority of patients had early-stage disease (183 stage I-IIIA), but 9 patients had oligometastatic disease at diagnosis and still received surgery of the primary tumor. All patients had provided written informed consent at the time of sample collection for genomic analysis. The SPECTAlung study was approved by several ethical committees (Ethische Commissie Onderzoek UZ/KU Leuven, in Belgium (S57513), Comité de protection des personnes “Ile-de-France VII”, in France (15-030 (PP 15-001))).

### Sample collection and processing

For each patient, H&E images were inspected by a pathologist to ensure a minimum of ten percent of viable tumor cells and to determine the amount of FFPE sections needed for DNA and RNA extraction. Additionally, from 120 patients (60 male, 60 female) from breast cancer, renal cell carcinoma, colorectal cancer, squamous cell head and neck cancer and NSCLC germline DNA was extracted from whole blood for sequencing to generate a “panel of normal” for mutation calling since for the NSCLC cohort investigated here, no matched normal samples were available.

### mIF data collection and processing with IFQuant

The mIF data for this project was in principle generated as described by Eling, Dorier et al^27^. Briefly, 3 sections of 4 µm thickness were used to generate mIF data with three antibody panels (**Supplementary Table 2**). The first section for mIF was followed by two sections for IMC and then the second and third section for mIF. Briefly, an automated Ventana Discovery Ultra staining module (ROCHE) was used for staining using primary antibodies, HRP-labeled antibodies, and OPAL reactive fluorophore detection. Imaging was performed using a PhenoImager HT (Akoya) and 8-bit QPTIFF images exported.

IFQuant was used to process mIF data as described in Eling, Dorier et al^27^. Briefly, channel unmixing was performed followed by nuclear segmentation and cell segmentation by expansion of up to 5 µm. For each cell different statistics of per cell fluorescence were recorded and thresholds for marker positivity applied to detect marker positivity. B cell patches were detected based on CD20 positivity if at least 40 B cells had a local density of 2000 cells/mm^2^ or more. Surrounding healthy tissue was annotated by a pathologist. For each patient sample TSV files containing the fluorescence intensity values per channel, X and Y location and marker positivity were exported.

### mIF data analysis

To identify cell types per mIF panel three phenotype keys were used that link the marker positivity to a phenotype (**Supplementary Table 2**, https://github.com/ImmucanWP7/mIF_cell_types). For each sample, a convex hull was generated around the tumor tissue based on the pathologist annotations and the area of each tumor was extracted and used to normalize cell type counts to generate cell type densities per mm^2^. Total abundances of cells were calculated as the sum of individual cell types within the tumor region. Cell-cell interaction graphs were generated using imcRtools. We used expansion from cell centroids and a distance parameter of 17 µm, corresponding to 4 µm distance between cells on average (average cell radius 6.8 µm). Cell-cell interactions were calculated with imcRtools utilizing the generated expansion graphs. For each cell type pair (e.g. A and B) directional interactions were calculated as the sum of interactions among A and B divided by either the abundance of A (average interactions of A and B for cells of type A) or the abundance of B (average interactions of A and B for cells of type B).

### Antibody conjugation and antibody mix preparation

Antibody conjugation and panel generation were performed as described in Eling et al. Briefly, antibody conjugation was performed following the manufacturer’s protocol using the MaxPar labelling kits (Standard Biotools). Conjugated antibodies were stored at 500 µm/mL in tris-based antibody stabilizing solution (Candor Bioscieces) at 4 °C.

### IMC1 staining and data collection

The IMC data for this project was collected under IMMUcan^26^ specifications. Details and a discussion about the IMMUcan workflow can be read in Eling et al.^27^. Briefly, FFPE slides were processed in batches of 19 slides. Each batch was removed from –80 °C, equilibrated to room temperature, deparaffinized, rehydrated, and stored in TBS (pH 7.6). After blocking, slides were stained with IMC1 (Supplementary Table 2) at 4 °C over-night using aliquots of ready-to-use antibody mixes stored at −80°C supplemented with fluorescent-labeled antibodies for region selection: pan Cytokeratin-Alexa-Fluor-488 (Thermo Fisher Scientific # 53-9003-82), CD45-DyLight-550 (Novus Biotechnologicals #NBP2-34528R), and CD163-Alexa-Fluor-647 (Abcam #ab218294). After washing with TBS and nuclear staining with Iridium intercalator (1 µM) and Hoechst solution (2 µg/mL) slides were washed again, dipped into ddH2O and dried under air flow.

The dried slides after staining were scanned within 48 hours with a Zeiss AxioScan with 10x magnification and filters for DAPI, 488, 555 and 647 using the ZEN software. Subsequently, Panoramas were generated for the batch of slides using a robotic slides loader for Hyperion+ (Standard BioTools) with customized software. The czi files from the slide scanner and the Panoramas (MCD files) were aligned using a custom napari plugin. After landmark-based alignment, three regions of interest (ROIs) for a total of ∼1 mm^2^ were selected to contain 30-70% or tumor cells together with immune and stromal cells^27^. Affine transformation was used to obtain the coordinates of the selected ROIs in mIF space in IMC space which were exported for acquisition on the Hyperion+ in batch mode using the robotic slides loader with 400 Hz and 4 dB ablation.

### IMC2 staining and data collection

Consecutive tissue sections to IMC1 were stained using a Leica Bond autostainer with IMC2 (**Supplementary Table 2**). After staining, brightfield scans of prominent tissue areas (partial Panoramas) were generated using the Hyperion+ Imaging System (Standard BioTools) with a robotic arm (Meca500, Mecademic) for automatic slide loading^27^. To identify regions of interest (ROIs) for IMC2, the IF images from the consecutive slide for IMC1 were aligned with the partial panoramas from the section for IMC2 via manual landmark-based alignment using immucan-roi^27^. Once registered, ROIs were selected to contain 30-70% of tumor cells mixed with immune and stromal cells and that did not overlap with ROIs for IMC1. Of note, we reasoned that due to rotation and missing alignment precision we would not be able to match ROIs across IMC1 and IMC2 without acquiring larger tissue areas which was not possible given time constraints of IMMUcan. We also reasoned that given the overlap in cell types and states across IMC1 and IMC2 measuring additional regions would be beneficial over measuring matched regions. After selection, the ROI coordinates were exported and acquired in batch mode with 400 Hz and 4 dB ablation.

### IMC data preprocessing

The steinbock toolkit^58^ (v0.14.1) was used to process the MCD files to generate hot pixel filtered TIFF files. Segmentation was performed using the Mesmer implementation within steinbock. Histone H3 and Iridium were used for nuclei detection for IMC1 and IMC2 and E-cadherin, CD3, CD8, CD20 and CD163 were used as markers for cellular boundaries for IMC1 and pan CK, E-cadherin, CD20, CD3, CD8 for panel two. Cellular interactions were quantified as cells that touch after a cell boundary expansion of 4 μm. For each cell and per panel the mean marker intensities, cellular area, centroid, major and minor axis length and eccentricity was calculated for downstream analysis.

### IMC2 cell type classification

To assign cell types to single cells we used a random forest classifier for IMC1^27^ and a XGBoost classifier for IMC2. For IMC2 we labeled a total of 182,000 cells using cytomapper^59^. The labeled images were split into training (80%) and test (20%) data. To train a multi-class classifier, we used the XGBoost framework in R using the xgboost and caret packages with soft-probability outputs. A grid search was performed to identify the optimal hyperparameters, evaluating combinations of subsample (0.7, 0.9), colsample_bytree (0.8, 0.9), max_depth (4, 6, 8), and eta (0.1, 0.2). Each model was trained with a maximum of 1000 boosting rounds and early stopping after 10 rounds of no improvement based on multiclass error. To prevent overfitting, grouped 5-fold cross-validation was applied using sample-level grouping, ensuring that cells from the same sample ID were not simultaneously present in both training and validation folds. For each parameter set, the model’s performance was evaluated using confusion matrices, sensitivity, specificity, and detection rate across classes. Of note, in IMC2 we did not include an antibody against CD4 to identify CD4 T cells. Instead, all CD8^-^FoxP3^-^ T cells were named CD4 T cells. In IMC1 neutrophils were defined as CD15^+^ MPO^+^ while in IMC2 we distinguished between PMN-MDSCs (CD15^+^CD66b^+^MMP9^+^/Arg1^+^), MDSCs (CD11b^+^CD15^-^CD66b^-^MMP9^+^/Arg1^+^) and neutrophils (CD15^+^CD66b^+^ Arg1^-^/MMP9^-^).

### IMC cell subtype definition

To identify fine-grained cell subtypes across IMC1, and to match them with the mIF panel 2 which contained PD1, GrzB, Ki67 and PD-L1, we defined cohort-wide positivity thresholds for CD8, GrzB, Ki67, PD1, PD-L1 and TCF7. To identify T cell subtypes, we grouped BnT cells into CD8 or CD8^-^. For CD8 and BnT_CD8 cells we then defined the following CD8 T cell sub-types based on positivity: PD1^+^Ki67^+^GrzB^+^TCF7^+^/^-^, PD1^+^Ki67^+^GrzB^-^TCF7^-^, PD1^+^Ki67^-^GrzB^-^TCF7^-^, Ki67^+^GrzB^+^PD1^-^TCF7^+^/^-^, Ki67^+^PD1^-^Grzb^-^TCF7^-^, GrzB^+^TCF7^+^PD1^-^GrzB^-^, TCF7^+^Ki67^-^PD1^-^GrzB^-^, GRZB^+^Ki67^-^PD1^-^TCF7^-^, Ki67^+^PD1^-^GrzB^-^TCF7^-^, PD1^+^GrzB^+^Ki67^-^TCF7^-^, PD1^+^TCF7^+^Ki67^-^GrzB^-^.

Similarly, for BnT_CD8^-^ and CD4 T cells based on PD1 and TCF7 we defined PD1^+^TCF7^+^, PD1^+^TCF7^-^ and TCF7^+^PD1^-^ CD4 T cell sub-types. Using the PD-L1 threshold we defined PD-L1^+^ macrophages and DCs. To identify sub-types of tumor cells, we clustered tumor cells with HLA-DR, CarbonicAnhydrase, cleavedPARP, PD-L1, IDO1, CD15, Ki67 and B2M markers using the R-phenograph implementation with 40 neighbors and subsequently meta clustered using kmeans clustering.

For IMC2 we used graph-based clustering for CD4 and CD8 T cells to identify cell subtypes using only T cell markers (CD7, Bcl2, GATA3, PD1, NKG2A, CXCL13, EOMES, CD137, CD134, Tbet, GITR, Tim3, Ki67). For graph-based clustering, different values for k (30-80), edge weighting (rank, jaccard) were tested with Louvain commutiy detection using the clsuterSweep function from the bluster package. Silhouette width and neigbhorhood purity were inspected. For CD4 and CD8 T cells we selected k = 70 and rank-based edge weighting. Inspection and clustering (euclidean distances and ward.D2 clustering) of the mean expression of markers across all cells from each cluster revealed 3 clusters for CD4 T cells and 2 clusters from CD8 T cells that showed no interpretable expression and where therefore merged. All clusters were given names based on the expression of markers. To identify activated Tregs we applied gaussian mixture models from the mclust and mixtools packages to identify positivity thresholds for CD137. Both models suggested an arcsinh transformed expression of 0.34 and 0.35 as cut off. We chose a more conservative cutoff of 0.5 to identify CD137 positive Tregs.

### Cellohood spatial cluster generation

Spatial clusters were generated using the Cellohood model as described in Możejko *et al.* (SUBMITTED). Briefly, Cellohood models *cellular neighborhoods*—spatially coherent groups of cells within tissues—by first hierarchically clustering spatially adjacent cells using a 25 µm distance threshold to form cell bags, each representing a local microenvironment composed of nearby cells with measured molecular profiles. A permutation-invariant set-transformer autoencoder was trained to encode each bag’s full expression matrix into fixed-size bag embeddings, preserving both per-cell information and collective spatial context. Permutation invariance is essential because each cell bag is modeled as a set, which lacks any inherent ordering of its elements. The model is trained to reconstruct the original marker data at the single-cell level using a mean-squared reconstruction loss and a permutation-matching module that aligns reconstructed and input cells—ensuring that the reconstruction loss can be computed meaningfully despite the unordered nature of the input. In this work, the analysis was performed at the bag level, meaning that each embedding represented the molecular and spatial characteristics of an entire local neighborhood rather than individual cells. To explore tissue organization at multiple levels of detail, Cellohood applies Principal Latent Dimension Analysis (PLDA) to control embedding resolution—from coarse to full representations. Here, we used the full-resolution (PLDA.100) embeddings, which captured the complete latent variation in the data and enabled identification of 36 distinct CNs representing the full spectrum of spatial phenotypes in the analyzed tissue. For per patient summaries, the counts of cells belonging to each type of CN per patient was used for differential abundance testing or the area normalized density of the count was used for CN densities.

### IMC data analysis

To investigate the expression of markers across cell types we calculated the mean expression of cell type specific markers per panel and cell type. Our list of cell type relevant markers can be found in **Supplementary Table 2**. Wilcoxon rank sum tests were used to estimate the difference in expression of markers on cell types per condition and FDR correction using Benjamini Hochberg was applied per condition. Cell-cell interactions were calculated as for mIF except that we utilized the steinbock generated interaction graphs (max 4 µm distance for interaction). All IMC ROIs were taken from within the tissue and therefore cellular densities were calculated as the sum of cell per patient divided by the area of the respective images. Cellular abundances were generated per patient.

### Differential abundance testing

For each data type (IMC1, IMC2, mIF1-3, spatial clusters IMC1 and IMC2) the absolute abundances per patient were used as input. Cell type abundances were analyzed using negative binomial generalized linear models implemented in edgeR (v3.40.2). Dispersions were estimated using all patientswithout a mean–variance trend, and robust model fitting was used to account for outliers. Differential abundance between groups was assessed using likelihood ratio tests applied to model coefficients. FDR correction using Benjamini Hochberg was performed within each comparison (e.g. *EGFR*-mutated vs *EGFR* wild-type).

### Infiltration scores

To estimate the infiltration of CD8 T cells into tumors we utilized the interaction counts between CD8 T and tumor cells. CD8 T cells with up to 2 interactions with tumors were classified as “bystanding” and those with more than two interactions with tumor cells were classified as “infiltrating”. The infiltration ratio was calculated as the ratio of infiltrating/bystanding for IMC1, IMC2 and mIF panel three. A final infiltration ratio per patient was calculated as the mean infiltration across modalities.

### RNA and WES data generation

DNA and RNA from FFPE and Frozen tissue were isolated using AllPrep DNA/RNA FFPE kit (Qiagen) or MagMAX FFPE DNA/RNA Ultra Kit (ThermoFisher). Exome libraries were generated using the Twist Human Core Exome Kit supplemented with the RefSeq and Mitochondrial Panel for enrichment (Twist Bioscience) or Kapa Hyperexome kit (Roche). Germline DNA for the generation of a “panel of normal” was extracted from EDTA tubes using PAXgene Blood DNA kit (Quiagen). Exome libraries were generated using the Twist Human Core Exome Kit supplemented with the RefSeq and Mitochondrial Panel for enrichment (Twist Bioscience) or Kapa Hyperexome kit (Roche). Tumor DNA was sequenced at a coverage of 100-150X or higher, gDNA at 30X or higher. Depending on the quality and quantity of the available RNA input material, the total RNA libraries were prepared using the KAPA RNA HyperPrep Kit with RiboErase (H/M/R) Globin (Roche) or the SMART-Seq Stranded Kit (Takara Bio). Next-generation sequencing was performed on an Illumina NovaSeq 6000 platform (Illumina) with 2 x 100 bp read mode. The generated sequencing data were demultiplexed using Illumina bcl2fastq (2.20) and adapters were trimmed with Skewer (version 0.2.2).

### RNA read mapping and count quantification

Paired-end sequencing reads were aligned using STAR (version 2.7.10b) against the GRCh38/hg38 human reference genome. Gene level counts were computed with LiBiNorm using the Gencode v29 assembly. Based on the RNA quality different library preparation kits had been used and therefore genes that were not expressed in one library type were removed. Furthermore, only genes with at least 1 cpm in at least 10% of samples were kept. Batch correction was then performed using ComBatSeq from the sva package and the resulting batch corrected counts were normalized using the calcNormFactors and cpm functions from the edgeR package. This data was used as input for pathway activity calculations. For MOFA, after batch correction, the variance stabilizing transformation from the DESeq package was applied for normalization.

### RNA Pathway activity

We used decoupleR^60^ on normalized (batch corrected, cpm) RNAseq counts using mlm method to quantify the activity of active signaling pathways described in PROGENy^61^ (Pathway RespOnsive GENes for activity inference). Retrieved pathway scores were used to compare groups of patients using t-tests. In particular, the 3 PROGENy pathways linked to inflammation (NFkB, MAPK and JAK-STAT) were tested for differences across the patient groups using stat_compare_means function in R.

### Cell type deconvolution from TCGA LUAD

Pre-calculated deconvoluted data for LUAD from TCGA was obtained from the Xcell^62^ website (https://comphealth.ucsf.edu/app/xcell) and clinical stages 1-3, treated as numeric values, were used to predict deconvoluted cell type estimates in order to investigate linear relationships between stage and cell type estimates.

### WES data processing

Whole exome sequencing data have been analyzed using the nextflow VEGAN pipeline (v2.1.1) available at https://github.com/bioinfo-pf-curie/vegan^63^. Briefly, sequencing reads have been aligned on the Human hg38 genome using Bwa-mem. Reads intersecting the exome capture with a minimum mapping quality of 20 were kept for downstream analysis. Duplicates were removed using the Picard MarkDuplicates tool. Somatic mutations have been called using a panel of normal, following GATK good practices with the Mutect2 software. Somatic variants annotation was performed with snpEff. Tumor Mutational Burden has been called using the pyTMB package^64^ included in the VEGAN pipeline. Only somatic variants with a VAF > 10%, a sequencing depth of > 20, and annotated as coding, non-synonymous variants were used. Polymorphisms (based on Gnomad and 1K genome projects) were discarded.

Because for this retrospective patient cohort no matched normal samples were available, a panel of normal was generated for mutation calling generally following these steps: https://gatk.broadinstitute.org/hc/en-us/articles/360035531132.

### Driver mutation scoring

From the tableReports generated with the vegan pipeline we selected 26 oncogenes or tumor suppressor genes^65^ for analysis. Only variants that passed quality filtering from the VEGAN pipeline, had a minimal variant allele frequency of 10%, had at least 3 reads covering the mutant allele, and were contained in the COSMIC database were kept. Variants were grouped to belong to one of the following functionally relevant alterations: single and multiple missense_variants, stop_gained, frameshift_variant, disruptive_inframe_deletion, conservative_inframe_deletion, and combinations thereof.

### Mutational signatures

Single-base substitution mutational catalogs from VCF files were generated using SigProfilerMatrixGeneratorR^66^. To account for the difference in genomic coverage between WES and WGS, we normalized the catalogs using trinucleotide-specific correction factors derived from the NGS panel used for WES sequencing. We applied refitting to lung-specific mutational signatures using the FitMS method from the signature.tools.lib R package that prioritizes common, organ-specific signatures (tier 2) while incorporating additional rare signatures (tier 1) when statistically supported^67^.

### MOFA+

Data preprocessing was tailored to each view. For the RNA view, we selected the top 1,000 most variable genes across all samples on batch corrected, variance stabilizing transformed counts.

For cell type densites, single cell marker expression, and cell type interaction views—where features are derived from combinations of cell types and can be highly redundant—we applied a correlation-based filtering strategy to reduce dimensionality while preserving biological signal. Specifically, we computed pairwise Pearson correlation matrices within each view, transformed these into distance matrices using 1 – abs(correlation), and performed hierarchical clustering using the hclust() function. Clusters were defined using cutree() function with a height threshold of 0.2, corresponding to an absolute correlation of ≥0.8 within each cluster (each feature in the cluster had a Pearson correlation of at least 0.8 to each of the other features). A representative feature was retained per cluster, while others were removed, allowing downstream interpretability of correlated features. Cellohood clusters and binary features—comprising driver mutations and dichotomized scores for TPS, CPS, and TMB—were included without further filtering.

Prior to MOFA2 (v.1.8.0) training, we log-transformed the density and Cellohood views to reduce right skew and compress dynamic range. This was followed by z-score standardization of the RNA, expression, interaction, and Cellohood views to center the features and normalize their variance. These preprocessing steps aligned with the assumptions of a Gaussian likelihood model preferred by MOFA and helped ensure that features contribute more equally to the variance decomposition. Additionally, they improved numerical stability during model fitting, speed up model training, and facilitated interpretability of the resulting factor weights^46^. The binary view, consisting of mutation and dichotomized clinical features, was not transformed and was modeled using a Bernoulli likelihood. MOFA was run with default settings, except for the number of latent factors, which was set to 6 because in preliminary runs LF6 explained only 12% of the variance, which informed our decision to limit the model to six LFs, as additional factors contributed progressively less to the overall variation.

### GSEA for weights

For each MOFA factor the RNA features were ranked by their weights and applied GSEA using the GSEA() function from the clusterProfiler package, with Gene Ontology Biological Process (GO:BP) gene sets from the C5 collection retrieved via the msigdbr package (species = Homo sapiens, category = “C5”, subcategory = “GO:BP”). Enrichment was assessed using standard scoring (scoreType = “std”) and FDR correction (pAdjustMethod = “BH”), retaining gene sets with 10–500 members. Only up to 10 of the most significantly enriched gene sets per factor were selected for visualization.

### TCGA validation

To validate the survival associations observed in our cohort, we downloaded bulk RNA-seq data from the TCGA LUAD and LUSC cohorts. For each MOFA factor, we identified the top 10 RNA features with the highest absolute weights and constructed factor-specific gene signatures consisting of features with either positive (enriched) or negative (depleted) weights. We then used the simpleScore() function from the singscore package to compute an enrichment score for each TCGA sample. This function calculates rank-based values of enriched and depleted genes separately, and reports the difference between the two scores as a final per-sample signature score. This approach yields one interpretable score per patient per factor, enabling survival analyses in independent datasets.

### General data analysis notes

R version 4.2.2. was used throughout with Bioconductor version 3.16. Statistical test were performed as indicated in the figure legends. Statistical tests were done on a per patient basis typically either averaging or aggregating measurements of multiple images per patient before statistical testing.

