## supplementary figures for "Comprehensive characterization of early-stage Non-Small Cell Lung cancers with multi-modal data integration"

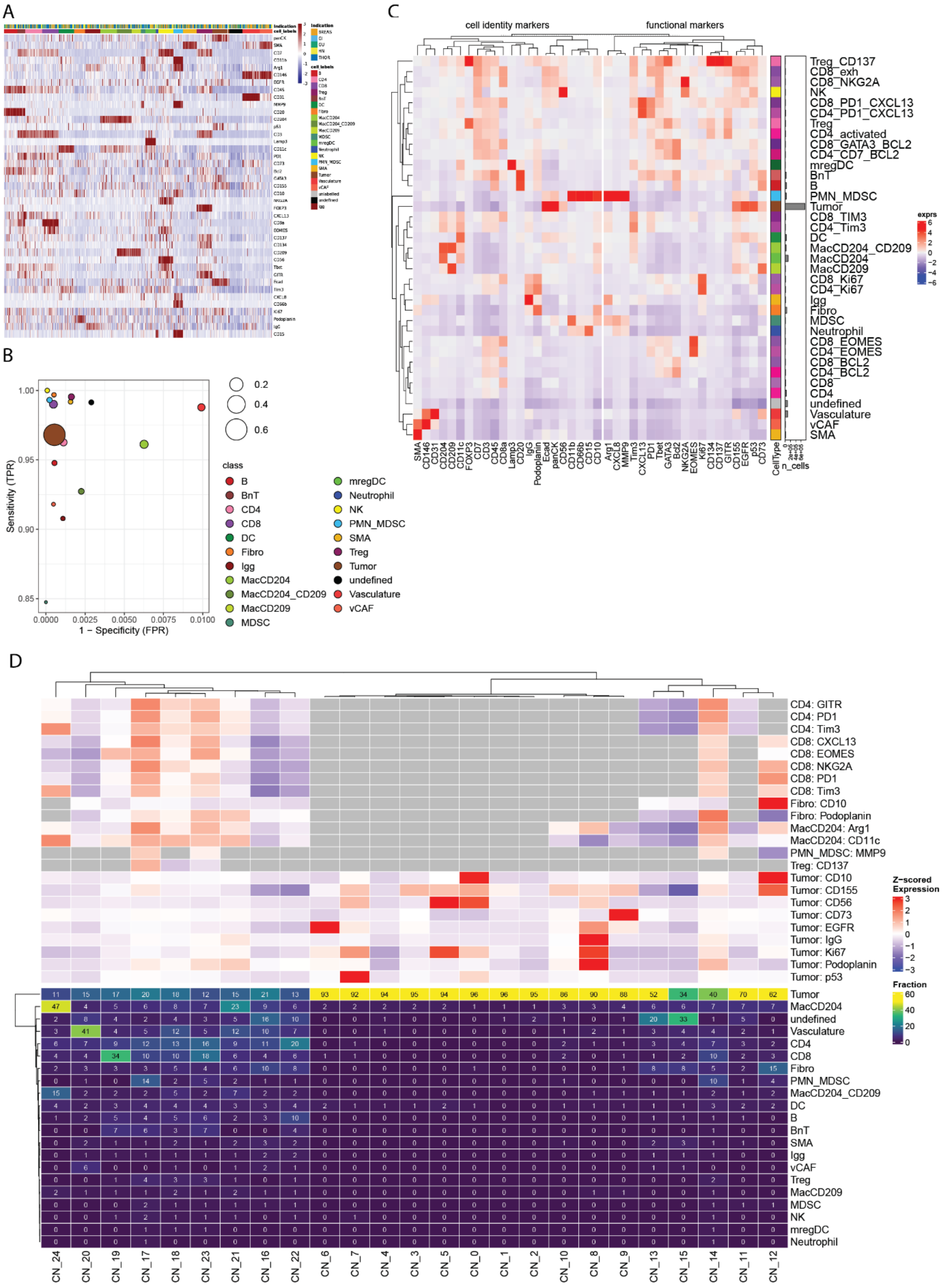


**Supplementary Figure 1: IMC2 cell typing, mIF correlation and spatial clusters**

**(A)** Heatmap depicting the z-scaled mean expression per image and cell type of the labelled data for cell classification. Top row indicates the cancer type and in the second row the color codes the cell type as shown in the legend. Classification was performed with labelled cells from five tumor indications as indicated in the legend: BREAS: Breast cancer; GI: Colorectal cancer; GU: Renal cell carcinoma; HN;

Squamous cell head and neck cancer; THOR: non-small cell lung cancer **(B)** Scatterplot indicating the true positive rates (TPR) and false positive rates (FPR) for the detection of individual cell types from IMC2 are shown based on a hold-out test data set. **(C)** Heatmap depicting the z-scaled mean expression of markers from IMC2 cell types and subtypes. Markers are split into cell type specific and functional markers. Bars indicate the abundance of cell subtypes across the cohort. **(D)** Heatmap depicting the fraction of cell types in each cellular neighborhood (bottom), and the mean z-scaled marker expression of cell type relevant markers in each cellular neighborhood are shown on top for IMC2. Mean marker expressions are shown for selected markers and only if the cell type made up at least 2 % or more of the spatial cluster.

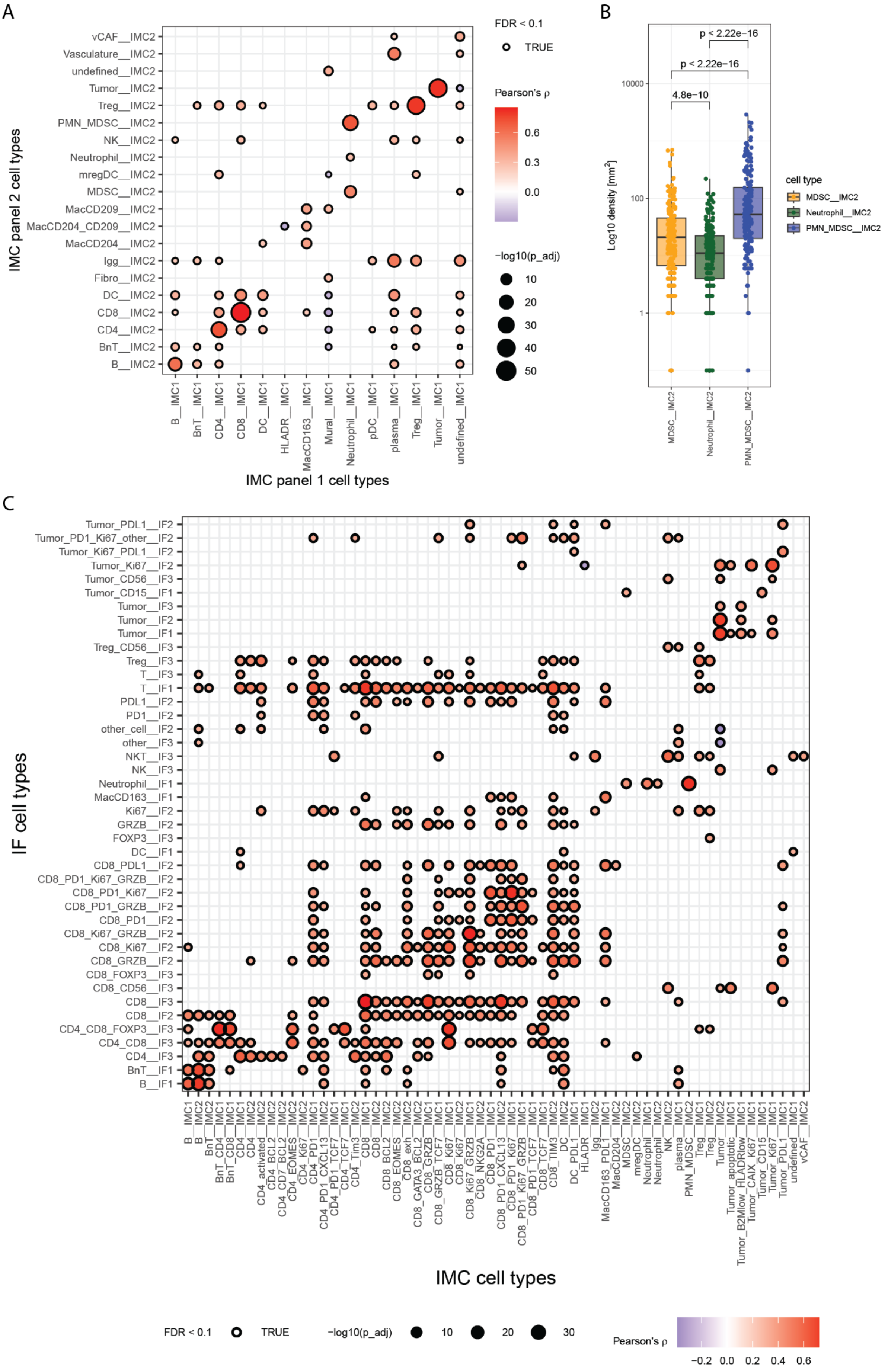

A
vCAF__IMC2
Vasculature__IMC2
undefined__IMC2
Tumor__IMC2
Treg__IMC2
PMN_MDSC__IMC2
NK__IMC2
Neutrophil__IMC2
mregDC__IMC2
MDSC__IMC2
MacCD209__IMC2
MacCD204_CD209__IMC2
MacCD204__IMC2
Igg__IMC2
Fibro__IMC2
DC__IMC2
CD8__IMC2
CD4__IMC2
BnT__IMC2
B__IMC2
IMC panel 2 cell types
B__IMC1
BnT__IMC1
CD4__IMC1
CD8__IMC1
DC__IMC1
HLADR__IMC1
MacCD163__IMC1
Mural__IMC1
Neutrophil__IMC1
pDC__IMC1
plasma__IMC1
Treg__IMC1
Tumor__IMC1
undefined__IMC1
IMC panel 1 cell types
FDR < 0.1
TRUE
Pearson's ρ
0.6
0.3
0.0
−log10(p_adj)
10
20
30
40
50
B
p < 2.22e−16
p < 2.22e−16
4.8e−10
10000
100
1
Log10 density [mm²]
MDSC__IMC2
Neutrophil__IMC2
PMN_MDSC__IMC2
cell type
MDSC__IMC2
Neutrophil__IMC2
PMN_MDSC__IMC2
C
Tumor_PDL1__IF2
Tumor_PD1_Ki67_other__IF2
Tumor_Ki67_PDL1__IF2
Tumor_Ki67__IF2
Tumor_CD56__IF3
Tumor_CD15__IF1
Tumor__IF3
Tumor__IF2
Tumor__IF1
Treg_CD56__IF3
Treg__IF3
T__IF3
T__IF1
PDL1__IF2
PD1__IF2
other_cell__IF2
other__IF3
NKT__IF3
NK__IF3
Neutrophil__IF1
MacCD163__IF1
Ki67__IF2
GRZB__IF2
FOXP3__IF3
DC__IF1
CD8_PDL1__IF2
CD8_PD1_Ki67_GRZB__IF2
CD8_PD1_Ki67__IF2
CD8_PD1_GRZB__IF2
CD8_PD1__IF2
CD8_Ki67_GRZB__IF2
CD8_Ki67__IF2
CD8_GRZB__IF2
CD8_FOXP3__IF3
CD8_CD56__IF3
CD8__IF3
CD8__IF2
CD4_CD8_FOXP3__IF3
CD4_CD8__IF3
CD4__IF3
BnT__IF1
B__IF1
IF cell types
B__IMC1
B__IMC2
BnT__IMC2
BnT_CD4__IMC1
BnT_CD8__IMC1
CD4__IMC1
CD4__IMC2
CD4_activated__IMC2
CD4_BCL2__IMC2
CD4_CD7_BCL2__IMC2
CD4_EOMES__IMC2
CD4_Ki67__IMC2
CD4_PD1__IMC1
CD4_PD1_CXCL13__IMC2
CD4_PD1_TCF7__IMC1
CD4_TCF7__IMC1
CD4_Tim3__IMC2
CD8__IMC1
CD8__IMC2
CD8_BCL2__IMC2
CD8_EOMES__IMC2
CD8_exh__IMC2
CD8_GATA3_BCL2__IMC2
CD8_GRZB__IMC1
CD8_GRZB_TCF7__IMC1
CD8_Ki67__IMC1
CD8_Ki67__IMC2
CD8_Ki67_GRZB__IMC1
CD8_NKG2A__IMC2
CD8_PD1__IMC1
CD8_PD1_CXCL13__IMC2
CD8_PD1_Ki67__IMC1
CD8_PD1_Ki67_GRZB__IMC1
CD8_PD1_TCF7__IMC1
CD8_TCF7__IMC1
CD8_TIM3__IMC2
DC__IMC2
DC_PDL1__IMC1
HLADR__IMC1
Igg__IMC2
MacCD163_PDL1__IMC1
MacCD204__IMC2
MDSC__IMC2
mregDC__IMC2
Neutrophil__IMC1
Neutrophil__IMC2
NK__IMC2
plasma__IMC1
PMN_MDSC__IMC2
Treg__IMC1
Treg__IMC2
Tumor__IMC2
Tumor_apoptotic__IMC1
Tumor_B2Mlow_HLADRlow__IMC1
Tumor_CAIX_Ki67__IMC1
Tumor_CD15__IMC1
Tumor_Ki67__IMC1
Tumor_PDL1__IMC1
undefined__IMC1
vCAF__IMC2
IMC cell types
FDR < 0.1
TRUE
−log10(p_adj)
10
20
30
Pearson's ρ
−0.2
0.0
0.2
0.4
0.6

**Supplementary Figure 2: Correlation between IMC and mIF data.**

**(A)** Pearson correlation of cell type densities from IMC_p1 (x-axis) and IMC_p2 (y-axis). For better visibility only correlations larger 0.2 or smaller -0.2 are shown color coded. The size of each point corresponds to the Benjamini Hochberg adjusted p-value and the black circle indicates values below 0.1. **(B)** Boxplot depicting the density of neutrophils, MDSCs and PMD-MDSCs (y-axis) across the dataset. Results from Wilcoxon from pairwise Wilcoxon rank sum tests are shown for all comparisons. **(C)** Pearson correlation of cell type densities from IMC (x-axis) and mIF (y-axis). For better visibility only correlations larger 0.3 or smaller -0.3 are shown color coded. The size of each point corresponds to the Benjamini Hochberg adjusted p-value and the black circle indicates values below 0.1.

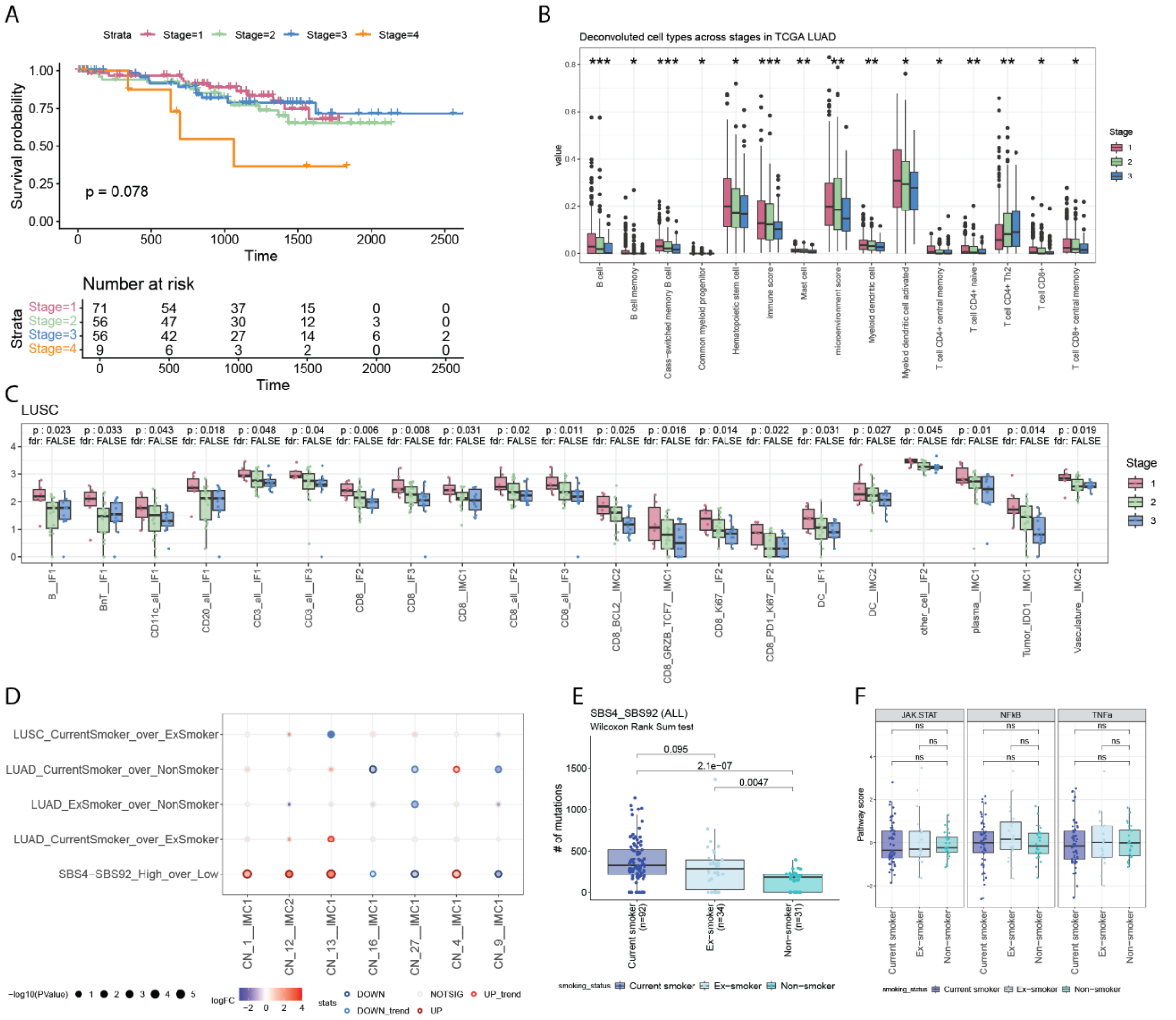

**Supplementary Figure 3: TME differences across clinical stages**

**(A)** Kaplan Meier plots for overall survival for clinical stages I-IV. **(B)** Boxplots depicting the densities of deconvoluted bulk RNAseq data from TCGA in LUAD across clinical stage. Statistics from linear models using numeric values ranging from 1 to 3 for stage as predictor for deconvoluted cell type values. Only results with an FDR < 0.1 are shown and p-values are coded as follows: p-value < 0.001 = ***, p-value < 0.01 = ** and p-value > 0.01 = *. **(C)** Same as in **(B)** using mIF and IMC cell subtype densities. Only results from models with p < 0.05 are shown and FDR < 0.1 is indicated with true/false. **(D)** Bubble plot indicating the differential abundances of spatial clusters across smoking categories in LUAD and LUSC separately. The size of each point corresponds to the p-value, the color to the log fold change and points are encircled to reflect statistical test and direction of effect (dark red: up regulation and FDR < 0.1; red: up regulation and p-value < 0.05; dark blue: down regulation and FDR <0.1; light blue: down regulation and p-value < 0.05). Spatial clusters for which at least one test (column) was significant are shown and FDR correction was performed within each categorical test (row). **(E)** Boxplot showing the number of SBS4 and SB92 signature associated mutations across smokers, former smokers and never smokers. **(F)** Boxplots depicting the differences in pathway activity scores (**Methods**) of JAK-STAT, NF-κB and TNF-α signaling for each smoking category. P-values were calculated using t-tests.

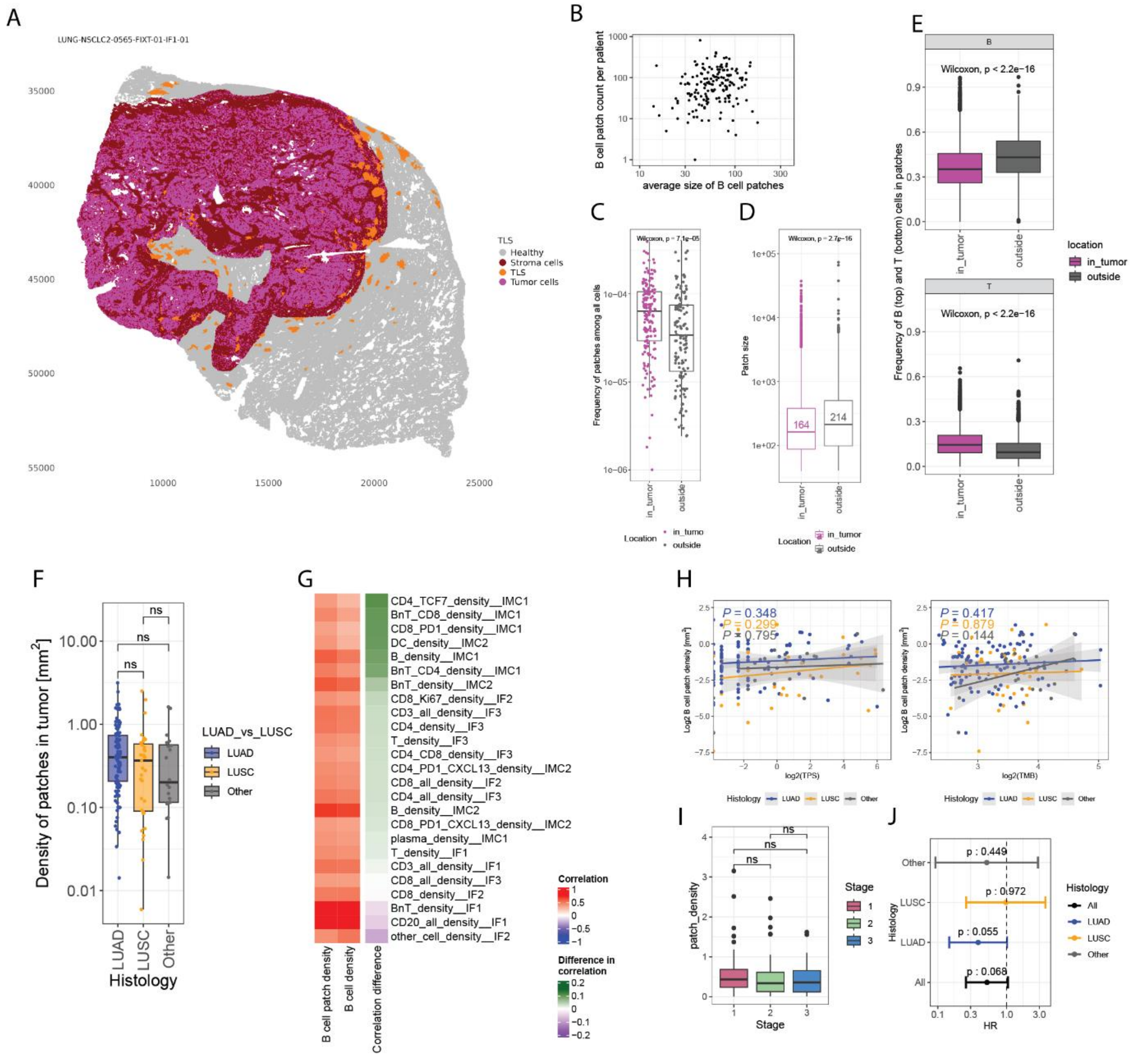


**Supplementary Figure 4: B cell patche across LUAD and LUSC**

**(A)** Schematic image of a tumor with each cell corresponding to a point colored by pink: tumor cells, red: stromal cells within the tumor, gray: cells annotated as belonging to healthy tissue, orange: cells belonging to a B cell patch. X and Y axis values represent distances in µm. **(B)** Scatterplot showing the average size of B cell patches per (x-axis) and the count of B cell patches (y-axis) per patient sample. **(C)** Boxplot showing the frequency of B cell patches when inside the tumor (+ 500 µm of the annotation) and the surrounding healthy normal tissue per patient. **(D)** Boxplot showing the average size of B cell patches when inside the tumor (+ 500 µm of the annotation) or within the surrounding healthy tissue. The median value is printed in red. **(E)** Boxplots showing the fraction of B cells (top) and T cells (bottom) within B cell patches when patches are located within the tumor (+ 500 µm of the annotation) or within the surrounding healthy tissue. **(F)** Boxplot showing the density of B cells patches per $mm^2$ for LUAD, LUSC and all other histologies. **(G)** Heatmap showing the spearman correlation of B cell patch densities (first column) and B cell densities (second column) with cell type densities (rows). The third column indicates the difference in correlation observed between B cell patches B cell densities. Rows are sorted based on the observed differences. **(H)** Scatterplots showing log2 B cell patch densities on the y-axis and log2 TPS values on the x-axis (left plot) and log2 TMB values on the x-axis (right plot). Points are colored by histology and p-values from linear models are shown per histology. **(I)** Boxplots showing the B cell patch densities across clinical stages. **(J)** Results from Cox proportional hazards models for groups of patients with high or low B cell patch densities (median split) are shown for LUAD (blue) and

LUSC (orange), other (gray) and all (black). P-values are indicated in the plot. Wilcoxon rank sum tests were performed for C, D, E, F, and I.

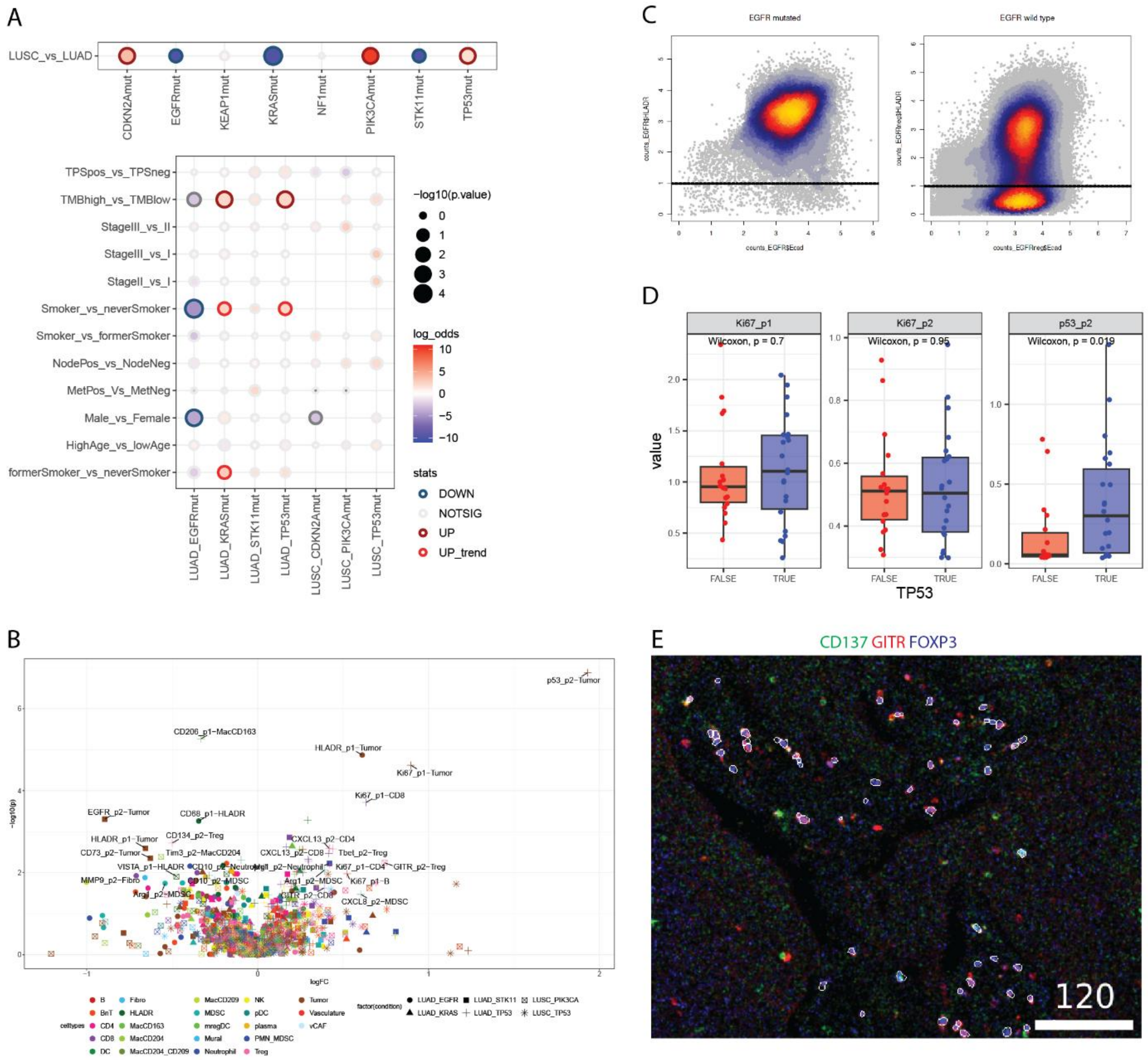

**Supplementary Figure 5: Driver mutations influence the TME**

**(A)** Bubble plot indicating the enrichment of mutations within histology (top) and other clinical parameters separated by histology (bottom) using Fisher tests. Points are colored by odds ratios and positive values indicate enrichment in the first string of the name (e.g. LUSC_over_LUAD: red indicates upregulation in LUSC). The size of each point corresponds to the p-value and points are encircled to reflect statistical test and direction of effect (dark red: up regulation and FDR < 0.1; red: up regulation and p-value < 0.05; dark blue: down regulation and FDR <0.1; light blue: down regulation and p-value < 0.05).. **(B)** Volcano plot showing the log fold change of the mean marker expression per cell type (x-axis) and the p-value from Wilcoxon rank sum tests (y-axis) for driver or tumor suppressor gene mutated samples vs respective wild type samples. Data points are colored by cell type and the shape corresponds to the respective mutation. Differential marker expression with FDR < 0.1 and an absolute log2 fold change > 0.33 is highlighted with text in the plot in the following fashion: Marker_IMCpanel_celltype. **(C)** Heatscatter plots showing the expression of HLA-DR (y-axis) and E-cadherin (x-axis) on single cells for EGFR-mutated tumor cells (left plot) and EGFR wild-type tumor cells (right plot). Dashed lines represent the cut point derived from a gaussian mixture model from the EGFR wild type data. **(D)** Boxplots showing the average expression per patient on tumor cells of Ki67 (IMC1) on the left, Ki67 (IMC2) in the middle and TP53 on the right for *TP53* mutated and wild type samples. P-values from Wilcoxon rank sum tests are indicated in the plots. **(E)** Example image showing the expression of CD137 (green), GITR (red) and FOXP3 (blue) on Tregs (outlined in white). Scale bar is given in µm.

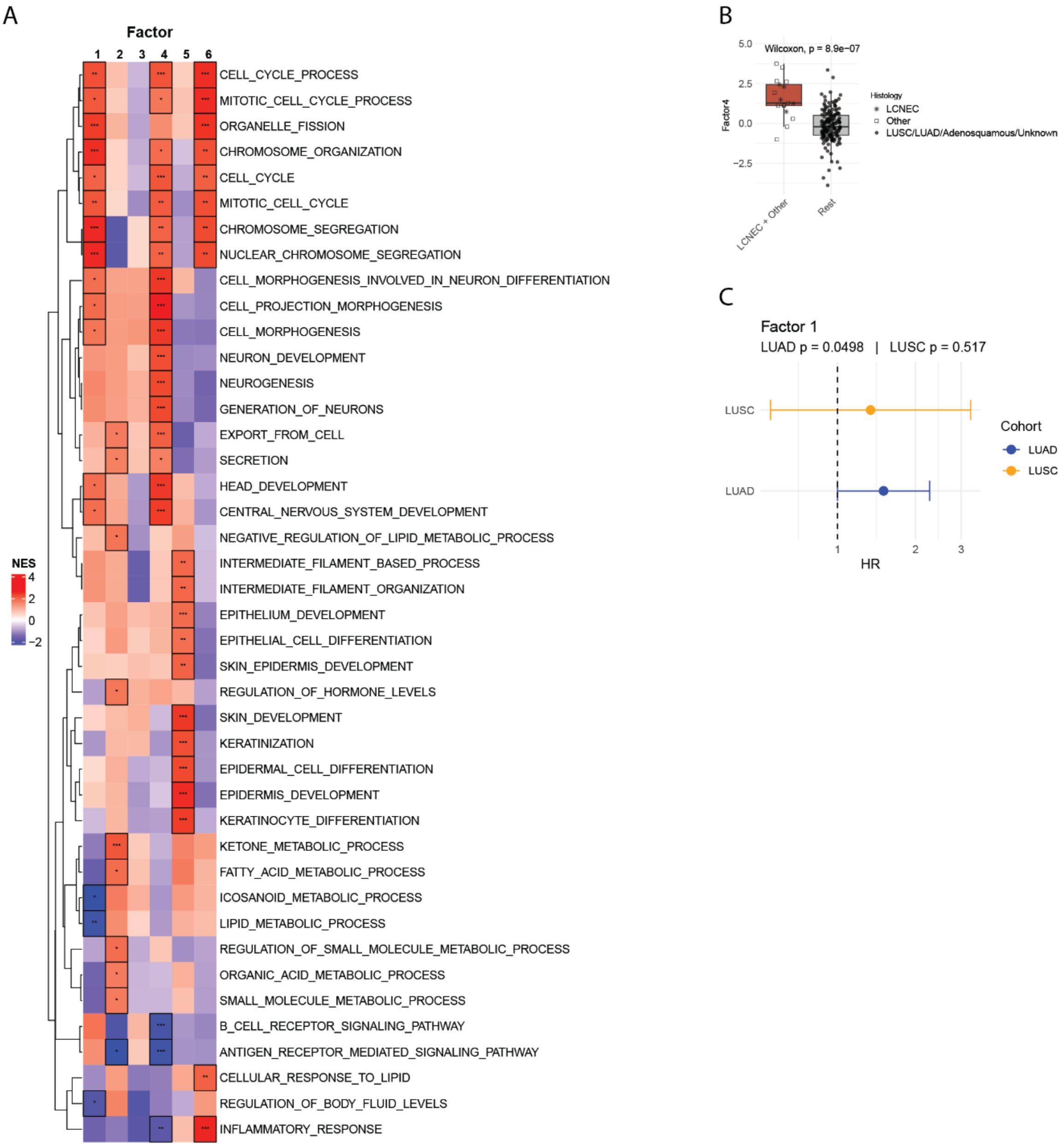


### Supplementary Figure 6: MOFA factor analysis

**(A)** Heatmap showing the normalized gene set enrichement scores across MOFA factors. Significant results are highlighted with black boxes and p-values are encoded as follows: p-value < 0.001 = ***; p-value < 0.01 = ** and p-value < 0.05 = *. **(B)** Boxplot comparing the values of LF5 across histologies. P-values were calculated using Wilcoxon rank sum test and are indicated in the plot. **(C)** Results from Cox proportional hazards models for continuous values of LF1 for LUAD (blue) and LUSC (orange) are shown.
